# Principles of gait encoding in the subthalamic nucleus of people with Parkinson’s disease

**DOI:** 10.1101/2022.02.08.22270370

**Authors:** Yohann Thenaisie, Kyuhwa Lee, Charlotte Moerman, Stefano Scafa, Andrea Gálvez, Elvira Pirondini, Morgane Burri, Jimmy Ravier, Alessandro Puiatti, Ettore Accolla, Benoit Wicki, André Zacharia, Mayte Castro Jiménez, Julien F. Bally, Grégoire Courtine, Jocelyne Bloch, Eduardo Martin Moraud

## Abstract

Disruption of subthalamic nucleus dynamics in Parkinson’s disease leads to impairments during walking. Here, we aimed to uncover the principles through which the subthalamic nucleus encodes functional and dysfunctional walking in people with Parkinson’s disease. We conceived a neurorobotic platform that allowed us to deconstruct the key components of walking under well-controlled conditions. We exploited this platform in 18 patients with Parkinson’s disease, which allowed us to demonstrate that the subthalamic nucleus encodes the initiation, termination, and vigor of leg muscle activation. We found that the same fundamental principles determine the encoding of walking. We translated this understanding into a machine-learning framework that decoded muscle activation, walking states, locomotor vigor, and freezing of gait. These results expose key principles through which subthalamic nucleus dynamics encode walking, opening the possibility to operate neuroprosthetic systems with these signals to improve walking in people with Parkinson’s disease.

**One Sentence Summary:** The subthalamic nucleus encodes the initiation, termination, and vigor of muscle activity, which supports real-time decoding of gait in people with Parkinson’s disease.

## INTRODUCTION

Every year, thousands of individuals with Parkinson’s disease (PD) undergo surgical implantation of electrodes in the subthalamic nucleus (STN). These electrodes not only enable the delivery of deep brain stimulation (DBS) to treat motor symptoms, but also allow recording of neuronal activity to study the encoding of functional and dysfunctional movements in the STN^1^.

Frequency domain analyses of local field potentials (LFP) combined with intraoperative recordings of single-cell activity uncovered fundamental principles through which the STN encodes upper-limb movements. It was found that the STN encodes the vigor of movements^2–5^, as well as the initiation, termination and concatenation of motor sequences^6,7^, which paralleled findings from animal models^8–12^. Moreover, abnormal modulation patterns in well-defined frequency bands of LFP signals have been shown to correlate with the severity of motor symptoms such as rigidity or bradykinesia^13,14^. This understanding has guided the development of closed-loop DBS protocols that maximize the treatment of these symptoms while reducing side-effects^15,16^.

Compared to upper-limb movements, gait and balance deficits respond insufficiently to pharmacotherapies and DBS interventions in the late-stage of PD^17^. Understanding the principles through which the STN encodes walking could support the development of treatments that alleviate these deficits. However, the few studies that investigated the encoding of walking in the STN reported conflicting results^18–21^.

Central to the understanding of upper-limb movement encoding was the possibility to study these principles under well-controlled experimental conditions restricted to single joints. Instead, walking involves complex sequences of dynamic movements requiring bilateral alternations between the right and left lower-limbs that are difficult to isolate and quantify. Due to this complexity, deciphering the encoding of walking in the STN has remained inconclusive.

To resolve this issue, we conceived a neurorobotic platform that emulated the key components of walking under well-controlled conditions, thus allowing us to isolate and quantify the encoding of lower-limb movements in the STN. We leveraged this platform to elucidate the principles that determine the encoding of leg muscle activations underlying walking. We validated these findings in the context of locomotor activities of daily living. Finally, we translated this understanding into machine learning decoders that detect functional and dysfunctional walking in realtime.

## RESULTS

### Neural recordings of subthalamic nucleus activity

We aimed to record LFP from the STN during locomotor functions. We included 18 patients with PD (**Table S1**) who exhibited severe motor fluctuations and varying deficits of gait and balance. The contribution of each participant to the different tasks is summarized in **Methods**.

Participants were implanted with DBS leads in the left and right STN (**Fig. 1A**), which were either externalized for five days after surgery (n = 8) or directly connected to an implantable pulse generator with sensing capabilities^22^ (Percept PC, Medtronic, USA) (n = 10). The location of DBS leads was confirmed using tridimensional anatomical reconstructions^23^ (**Fig. 1A** and **Fig. S1**).

**Figure 1.**
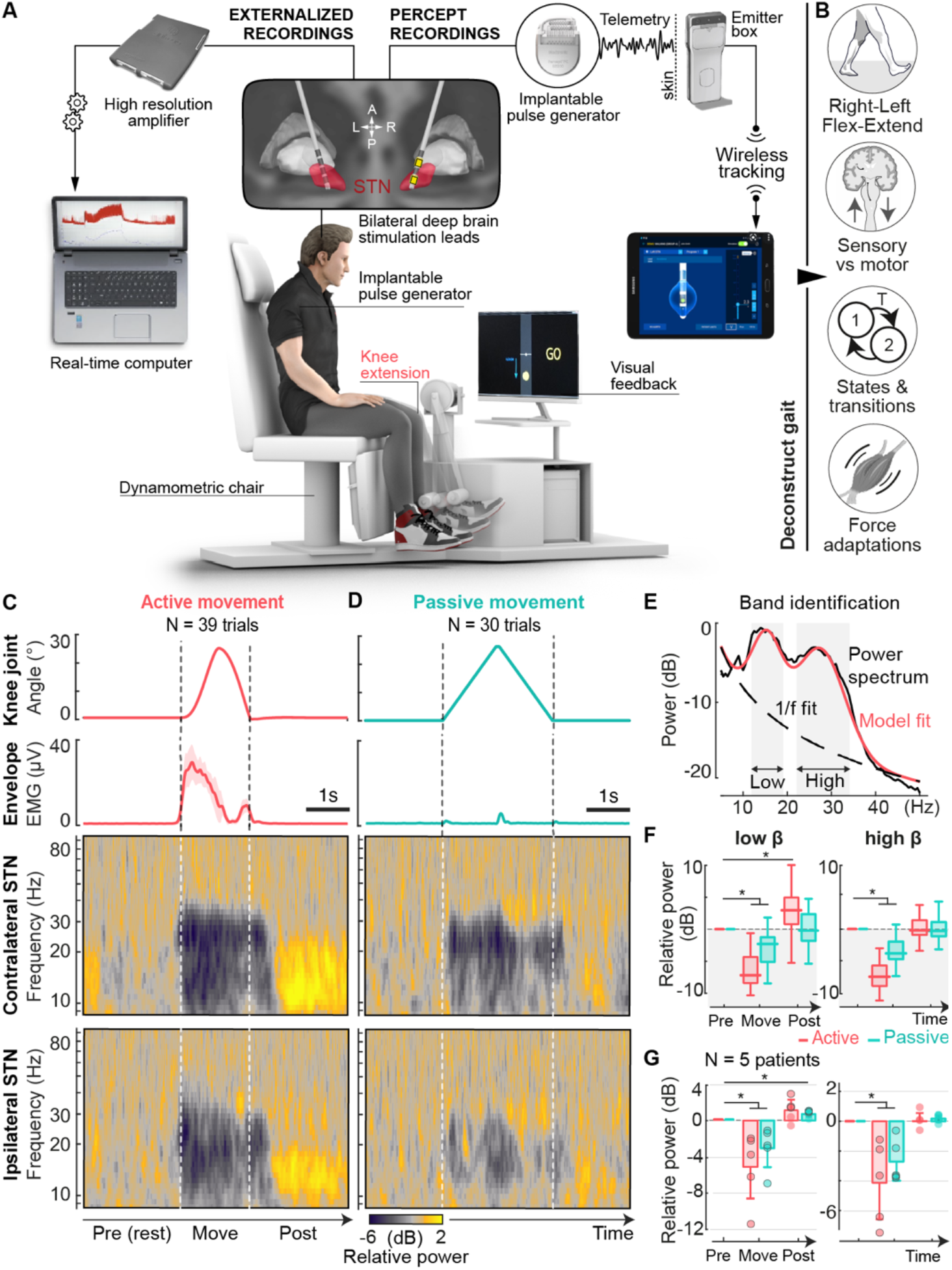
Bilateral STN modulations encode active and passive leg movements. **(A)** Neurorobotic platform to study the encoding of leg muscle activations in the STN in well-controlled conditions. Patients are sitting comfortably during unilateral knee-extension movements, the shin of their most affected leg attached to a rotating dynamometer. Movements are either passively or voluntarily performed. Bilateral electromyographic and STN signals are recorded concurrently to movement angular parameters. The location of DBS leads is verified through 3D anatomical reconstructions. Contacts used for LFP recordings (most affected hemisphere) are highlighted in yellow. **(B)** Deconstructed components of gait-related leg muscle activation to be studied in isolation. **(C)** Voluntary movement performed by participant P1. Movement angle (mean ±SD), knee extensor muscle envelope (vastus medialis, mean ±SD), and scalograms for contralateral and ipsilateral STN LFP normalized to baseline (rest pre-movement). Each movement epoch is preceded and followed by 2.5s of rest. **(D)** Same plots during passive movements of the knee joint. **(E)** Patient-specific band identification (same participant) using an unbiased fitting algorithm (see methods). Extracted low and high-beta bands are indicated as shaded boxes. **(F)** Boxplot of normalized power across trials (same patient) averaged over each window (rest pre, movement and rest post). **(G)** Group-average and standard deviation of baseline-normalized power of the low and high-beta bands of the contralateral STN. N = 5 participants performed this task (see methods). Dots represent the mean for each patient. * p<0.05 paired t-test with Bonferroni correction. Circled dots indicate significant differences from baseline for individual participants. The amplitude of modulations was more pronounced during active compared to passive movements (mean effect size (±SD) : low beta - 2.1dB±1.7dB, high beta -1.4dB±1.7dB, not significant t-tests).

We first assessed the electrophysiological characteristics of LFP from each pair of electrodes to identify the bipolar contacts with the highest beta power. To define patient-specific frequency bands, we employed an unbiased fitting algorithm that parametrized neural power spectra as a combination of an aperiodic (1/f) and several periodic oscillatory components24 (**Fig. 1E**). Low-beta activity was identified in the most affected STN in 11 participants, high-beta in 18 participants and low-gamma in 6 participants.

### Neurorobotic platform to study leg muscle activation

We aimed to conceive an experimental paradigm that mirrored the well-controlled conditions restricted to single joints that enabled elucidating principles of upper-limb movement encoding.

We established a neurorobotic platform to study leg movements and muscle activation patterns restricted to a single joint while patients were seated comfortably (**Fig. 1A**). Participants were secured in an instrumented dynamometric chair that enabled the concomitant recordings of leg kinematics, force, electromyographic activity, and bilateral STN LFP. A screen located in front of the participant displayed task instructions concomitantly to ongoing movement and force parameters (**Movie S1**).

We exploited this platform to design paradigms that deconstructed the key components of leg muscle activation underlying walking (**Fig. 1B**). Specifically, we designed paradigms that emulated the differences between (i) sensory feedback versus volitional leg muscle activation, (ii) transient versus sustained activation, (iii) extensor versus flexor activation, (iv) activation from differ-rent joints, (v) ipsilateral versus contralateral activation, and (vi) varying levels of muscle activation.

### Encoding of sensory feedback versus volitional leg muscle activation

We first recorded the patterns of STN modulation during self-paced knee-joint extension movements, which required the activation of quadriceps muscles (**Fig. 1C**). STN patterns displayed a stereotypical sequence of movement-related modulations, both contralateral and ipsilateral to the recruited muscles. These patterns involved a significant desynchronization during movement execution, both in low- and high-beta power, followed by a rebound after movement termination that predominantly occurred in the low-beta band (**Fig. 1F,G**).

This sequence of motor-related modulations suggested a direct link between STN activity and the volitional activation of leg muscles. However, walking generates a flow of sensory feedback that may also contribute to the modulation of STN LFP. To disentangle sensory feedback versus volitional leg muscle activation, we compared LFP modulations during active versus passive leg movements (**Fig. 1D**). Unexpectedly, we found that passive movements generated reproducible LFP modulations that mirrored the patterns underlying active movements. All tested participants had significant low- and high-beta desynchronization during passive movements, and three had a significant low-beta rebound after movement (**Fig. 1F,G)**. Nevertheless, the amplitude of these movement-related modulations was more pronounced during active compared to passive movements.

These results revealed that the STN encodes both volitional commands to activate leg muscles and sensory feedback associated with leg movements.

### Encoding of muscle activation dynamics

Walking involves transitions between different phases of gait, including transient activity of flexor muscles during swing, and sustained activation of extensor muscles during stance. Moreover, gait adjustments require volitional modulation of muscle activation levels. We designed paradigms that mimicked these conditions in the neuro-robotic platform.

We first asked participants to perform a transient, isometric knee-extension task with two levels of force (**Fig. 2A**). Despite the lack of movement, we found that the recruitment of the quadriceps muscles involved significant desynchronization and resynchronization profiles in low- and high-beta power that were similar to the patterns underlying active movements (**Fig. 2C**). Unexpectedly, these modulations were not influenced by the level of force production.

**Figure 2.**
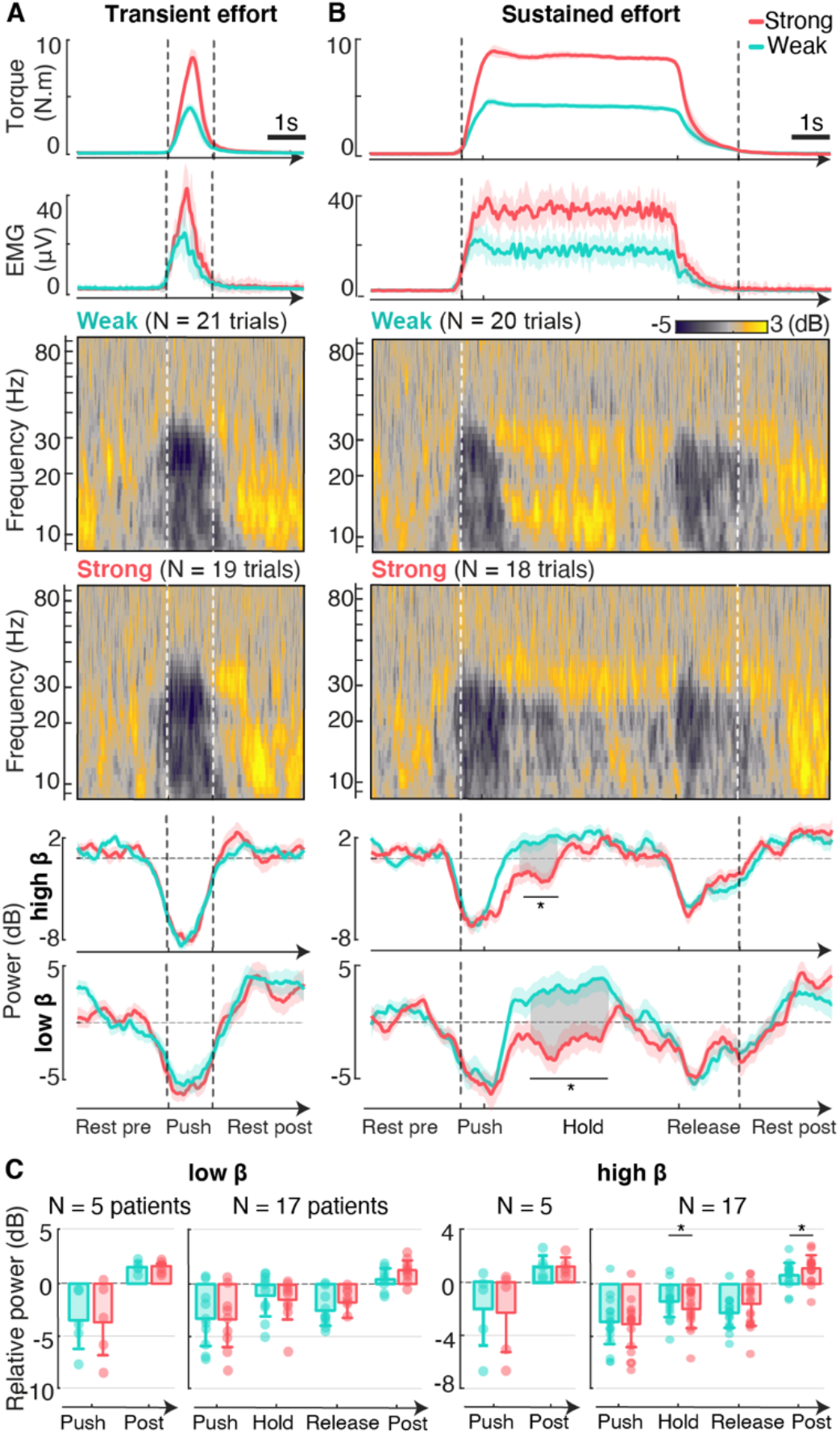
STN encodes muscle activation dynamics. Patients performed an isometric knee extension task with two levels of force (weak 33% of maximal voluntary contraction, strong 66%). **(A)** Task involving a transient effort (participant P1). Average traces of torque and knee extensor muscle envelope (mean ±SD), average spectrograms of the contralateral STN normalized to baseline (rest pre-movement, defined as 2.5s before the onset of effort) and traces of low- and high-beta power (mean ±SD). **(B)** Same plots for a task involving a sustained effort. Shadowed areas show significant differences in band-power when tasks performed with a weak and a strong force (Monte-Carlo cluster comparison). **(C)** Barplots of low- and high-beta power (mean ±SD) for both tasks. N = 17 patients performed the sustained task, only N = 5 of them performed the transient task. Low-beta was identified in all participants, and high-beta in 4 /5 and 10/17 participants respectively. * p<0.05 paired t-test with Bonferroni correction. No difference was found between levels of force for the transient task at the group level (two-way ANOVA, n=5: Effect of time window: df=2, low beta: F=2.64, p=0.09; high beta: F=8, p=0.002. Effect of force: df=1, low beta: F=0.04, p=0.83; high beta: F=0.02, p=0.90). Within participants: 5/5 (exhibiting low beta) and 4/5 (high beta) showed a significant effect of the time window (Two-way unbalanced ANOVA); 1/5 (low beta) and 0/5 had an effect of force. For the sustained task: 9/10 (low beta) and 17/17 (high beta) showed a significant effect of the time window. 3/10 (low-beta) and 7/17 (high-beta) had a significant effect of force.

We next designed paradigms that emulated the sustained activtion of muscles during the stance phase of gait. We asked participants to produce and maintain a constant activation of the quadriceps muscles with a weak or strong force (**Fig. 2B**). As observed during the transient task, the initiation of muscle activation led to a strong desynchronization in low- and high-beta power that was not modulated by the level of force produced.

In contrast, holding a sustained force required a continuous activation of muscles during which the amplitude of LFP modulations depended on the level of force. When holding a weak force, beta power rapidly resynchronized and stabilized around baseline. Instead, maintaining a strong force significantly delayedthe resynchronization of beta power (**Fig. 2C**). These patterns suggested that the STN reflects the vigor involved in the production of sustained leg muscle activation.

Termination of muscle activation also induced reproducible desynchronizations in low- and high-beta power that resembled the patterns underlying the initiation of the task (**Fig. 2B,C**). These modulations were not modulated by force at the group level, although they were more pronounced after weak than after strong efforts in some participants (p<0.05 for 4/10 participants exhibiting low-beta and 6/17 in high-beta). Post-effort rebounds in beta-power were also larger after strong than after weak force production.

These patterns of STN dynamics were similar during the concatenation of force levels (**Fig. S2**), across leg joints (knee vs ankle), and for extensor vs flexor muscles (**Fig. S3**).

### Bilateral encoding of muscle activation

Walking requires finely tuned coordination between left and right leg movements. We thus asked whether the activity of the STN incorporates information that is predominantly related to the contralateral versus ipsilateral leg during sustained muscle activations.

We analyzed bilateral recordings of the STN during the production of sustained levels of force from the left versus right leg (**Fig. S4)**. As expected, STN modulations were more pronounced when the movement was performed with the leg contralateral to the movement, although movement-related modulations systematically emerged in the ipsilateral STN as well.

### Idiosyncratic encoding of leg muscle activation

The neurorobotic platform allowed us to identify common principles underlying the encoding of the initiation, termination, and vigor of leg muscle activation in the STN, which were consistent for all participants. However, we also detected idiosyncrasies in the definition and dynamical behavior of frequency bands across participants (**Fig. S5** and **Fig. S6)**:

Our unbiased fitting algorithm extracted beta bands that differed across participants, including the presence of low-or high-beta, or a combination of both. The center frequency of these bands and their amplitude differed between patients. Moreover, some participants exhibited modulations in low-gamma power that also increased with the level of sustained force.

(i) Two participants displayed additional frequency bands that modulated with force. One participant (P10) exhibited modulations in the alpha-band. A second participant (E2) showed a synchronization around 20 Hz that scaled up with force^25,26^.

These differences stressed the importance of accounting for participant-specific features in the analysis and use of STN recordings.

### Real-time decoding of force production

The isolated conditions of the neurorobotic platform allowed to expose a link between STN modulations and the vigor of sustained leg muscle activation. This opened the intriguing possibility to predict the level of leg force production from STN dynamics.

We translated this hypothesis into a machine-learning framework that used Random Forest classification algorithms to decode leg force in real-time (**Fig. 3A**). Algorithms were trained to predict one of three classes (“rest”, “weak force” or “strong force”) based on the entire power spectrum of bilateral LFP modulations. Since vigor-related modulations in LFP occurred exclusively when muscle activations were sustained, we restricted the training of the classifiers to this specific period.

**Figure 3.**
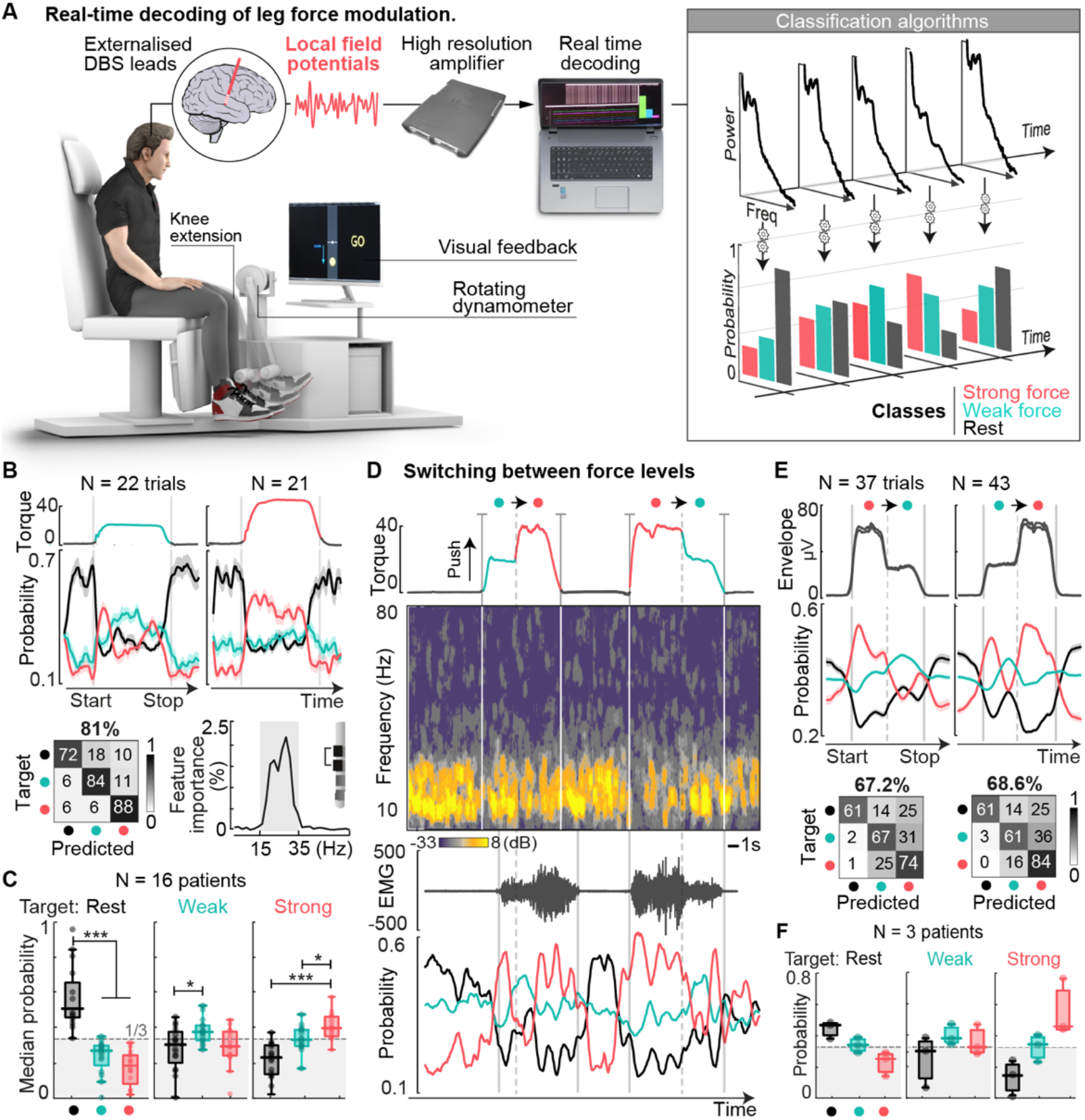
Real-time decoding of leg force modulation. **(A)** Experimental setup for real-time decoding experiments. Bilateral externalized DBS leads are connected to a high-resolution amplifier (sampling frequency 8kHz). Digitalized LFP signals are fed to a dedicated computer running real-time decoding algorithms that predict the probabilities of three classes (“rest”, “weak” and “strong” force) based on the spectral components of LFP over a moving window of 500ms. Algorithms were trained (and tested offline) on a first task, in which patients maintained a single force for 5s and then released the effort. Decoders were then tested in real-time on a second task, in which patients had to switch online from a strong force to a weak force, or vice-versa. **(B)** Decoder performance during the first force task (participant E1). Probability (median ±SEM) for each class over time, and confusion matrix (Accuracy = 81.06%, F-score = 0.79). Feature contribution traces display the frequencies that are automatically identified as being more relevant for the decoder (one contact from the contralateral lead shown). Relevant information is predominantly extracted from the low- and high-beta range for this patient (shaded box). **(C)** Average performance for all 16 patients tested on this task (one-way ANOVA with Bonferroni correction, n = 16, * p<0.05, * * * p<0.001) **(D)** Illustrative example of real-time recording during the second task, which involves switching between levels of force, for the same participant (E1). Two trials are shown, first a change from a weak force to a strong force, then a change from strong to weak. **(E)** Average performance traces (EMG envelope, top, and probability traces, bottom) and confusion matrices for both cases. **(F)** cross-patient performance achieved for the 3 patients tested in real-time.

We trained the classifiers offline for each participant. The algorithms automatically extracted patient-specific spectral features that captured their contribution to the encoding of muscle activation. These features closely matched the relevant frequency bands previously identified in each participant (**Fig. 3B**). The classifiers then combined these weighted features to discriminate rest from sustained weak-or strong-force conditions. Across the 14 participants, the classifiers predicted the three states with 67 ±10% SD cross-validation accuracy (**Fig. 3C** and **Fig. S8**), despite variability during the transitions between states.

We then evaluated the performance of these algorithms during real-time experiments. On the following day, participants performed a task during which they were required to alternate between weak versus strong levels of sustained force, based on instructions displayed on the screen (**Fig. 3D**). The classifiers acquired and processed LFP continuously, and automatically generated real-time predictions that closely matched the three motor states (average accuracy 68 ±11% SD) (**Fig. 3E,F** and **Movie S1**).

### Multimodal locomotor analysis platform

We sought to translate these principles into paradigms that enabled studying the encoding of leg muscle activity during walking.

We established a multimodal gait platform that allowed concurrent recordings of whole-body kinematics, bilateral leg muscle activity, and bilateral STN LFP during unconstrained walking (**Fig. 4A** and **Movie S2**). To repro-duce single-joint tasks with different levels of vigor, we instructed the participants to walk along a horizontal ladder with either short or long steps, which required weak or strong levels of muscle activation and force (**Fig 4B**).

**Figure 4.**
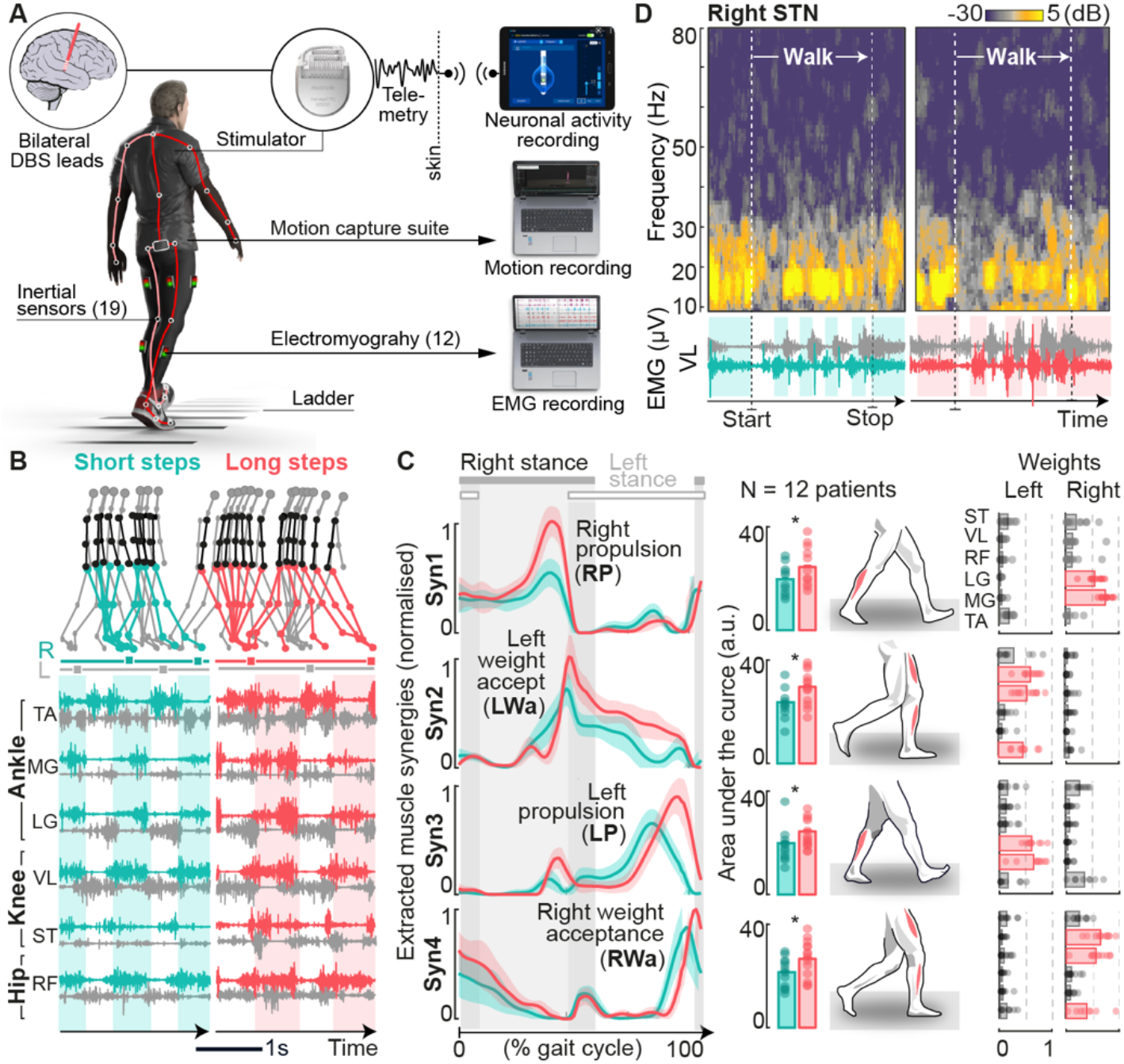
Multimodal locomotor analysis platform. **(A)** Experimental setup for monitoring motor and neural states during unconstrained locomotor tasks. Externalized patients wore a whole-body suit embedded with 19 inertial sensors located at key landmark joints of the body. This allowed to track their kinematics wirelessly and in real-time. All patients additionally wore six wireless electromyographic sensors on each leg that recorded agonist and antagonist muscles of the ankle (Tibialis Anterior TA, Medial Gastrocnemius GM, Lateral Gastrocnemius LG), knee (Vastus Lateralis VL, Semitendinosus ST) and hip (Rectus Femoris RF). **(B)** Patients performed various locomotor tasks requiring different levels of muscle recruitment and force. **(C)** Computation of leg muscle synergies. We extracted 4 muscle synergies from the set of 12 bilateral EMG signals across locomotor tasks, which accounted for up to 90% of the variance of the original data. The temporal profiles of these synergies over the gait-cycle (left) and the weights of muscle contributions across patients (right) indicated that the extracted synergies predominantly captured bilateral propulsion and weight acceptance phases. All 4 synergies exhibited a clear increase with force. * p<0.05 paired t-test. **(D)** Bilateral STN LFP were recorded concurrently and aligned in time with kinematic and electromyographic signals for each task. Visual inspection of raw spectrograms highlighted modulations in well-defined frequency bands over the course of the task and across tasks, which aligned to the beginning and execution of walking (participant E1).

Contrary to single-joint paradigms, walking involves the coordinated activation of multiple muscles from the right and left leg. To link STN modulations to changes across leg muscles, we synthesized the complex patterns of muscle activity during walking into lower-dimensional muscle synergies (**Fig. 4C** and **Fig. S9**). We computed synergies across all the recorded muscles from the left and right leg. We restricted synergy extraction to 4 components, which were sufficient to capture muscle activations underlying short versus long steps (∼90% of explained variance). The timing of these synergies suggested that they reflected propulsion and weight acceptance phases from the right and left leg.

### STN LFP encodes the onset and termination of walking

We previously found that the STN encodes the onset and termination of muscle activation. We thus asked whether the same principle emerges during walking.

We instructed participants to stand for approximately 3 seconds before initiating a sustained bout of walking, after which they were requested to stop and stand for another 3 seconds (**Fig. 5B**). As observed during single-joint tasks, the initiation of walking involved a robust desynchronization in low- and high-beta power. Likewise, a transient desynchronization occurred in the same bands during the step preceding the termination of walking (**Fig. 5C**).

**Figure 5.**
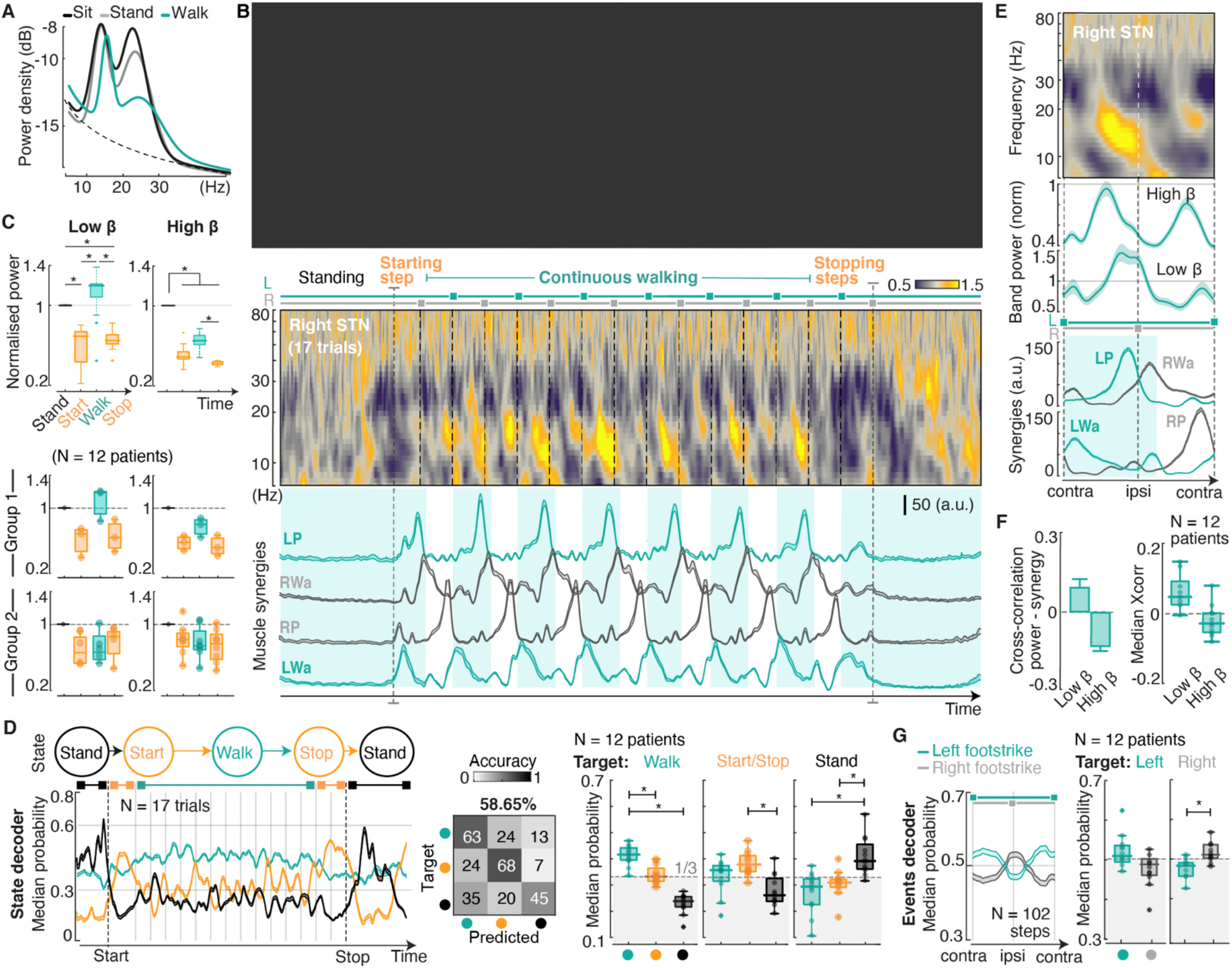
STN encodes transitions between walking states and modulations in leg muscle synergies. **(A)** Illustrative example of changes in power spectral density (participant P1). Walking induces an overall decrease of high-beta power as compared to sitting and standing, along with modulations in the amplitude and frequency range of low-beta power. **(B)** Time decomposition of a complete walking sequence for patient P1. Chronophotography (top, please contact the corresponding author to request access to the image), average spectrogram (middle) and synergy traces (median ±SEM) highlight modulations in low- and high-beta that exhibit well-defined temporal patterns during continuous walking, aligned to the activation of leg muscle synergies. Transitions between locomotor states (onset and termination of walking) exhibit strong desynchronizations across beta bands. **(C)** Quantification of median power in the low- and high-beta bands during standing, walking and state transitions for patient P1 (KruskalWallis test with Bonferroni correction, n=20). Across participants, we distinguished two groups: those for whom low-beta exhibited strong rebounds after effort, and those who had a tendency to remain desynchronized. **(D)** Automatic prediction of locomotor states. We trained decoding algorithms to automatically discriminate between 3 classes (“standing”, “walking” and “transitions”) based on bilateral STN LFP signals. Average probability traces for each class over time and confusion matrix for patient P1, and cross-patient performance (* p<0.05 one-way ANOVA with Bonferroni posthoc correction, n = 12) **(E)** Time decomposition per gait-cycle. Average spectrogram (top), low- and high-beta power traces (median ±SEM, middle) and synergy traces (bottom) aligned and interpolated between consecutive contralateral footstrikes. **(F)** Cross-correlation coefficients between band power (low- and high-beta) and synergy activations (sum of all four extracted synergies) for each gait-cycle. High-beta band is negatively correlated with synergy traces, and desynchronizes whenever muscles are recruited, whereas low-beta exhibits a positive correlation due to rebounds at the late stages of swing. **(G)** Automatic prediction of bilateral gait events. We adapted our decoding algorithms to discriminate right vs left foot strikes. Average traces aligned between contralateral footstrikes for participant P1, and cross-patient performance (* p<0.05, Signrank test, n= 12), show the capacity to discriminate between such events.

These results suggested that changes in the walking state of participants could be predicted from STN LFP. To test this idea, we trained our machine learning classification algorithms to discriminate the probability of three classes: “standing”, “start/ stop” transitions and “walking”. The temporal profiles of the predicted probabilities for each class and performance analyses confirmed that the classifier was able to predict these three states accurately (55 ±5% SD across participants) (**Fig. 5D, Fig. S11** and **Movie S3**).

### STN LFP encodes the timing of leg muscle synergies

STN modulations during single-joint motor tasks were irrespective of the type (flexor vs extensor), location (ankle vs knee), or side (ipsilateral vs contralateral) of the muscles recruited. We thus asked how the STN would encode muscle activation patterns during walking, since the cyclic patterns of leg movements require a succession of stance and swing phases from both legs, associated with a mixture of flexor and extensor muscle activations.

LFP analysis over the course of a gait cycle revealed modulations in low- and high-beta power that were locked to the activation of muscle synergies from both legs (**Fig 5E**). Concretely, high-beta power exhibited pronounced desynchronizations spanning mid-swing to early stance. This timing coincided with the concomitant activation of the propulsion muscle synergy from one leg and the weight acceptance muscle synergy from the other leg. These patterns occurred twice per gait cycle, as expected from the alternating movements of the left and right legs and bilateral encoding of leg movements in the STN (**Fig. S7**). Consequently, high-beta power traces correlated negatively with the activation of leg muscle synergies (**Fig. 5F**).

Modulation of LFPs were even more pronounced in the lowbeta band. Walking involved desynchronization patterns during the weight acceptance phase (double stance) and the transition from stance to swing, followed by a rebound during the ballistic part of swing, which also occurred twice per gait cycle. Low-beta power thus exhibited predominantly positive cross-correlations with leg muscle synergies (**Fig. 5F**).

These phase-locked patterns encouraged us to explore the possibility to predict gait events from STN LFP (**Fig 5E)**. The decoder was trained to predict the probability of left vs right foot strikes, and reached an accuracy of 60 ±8% SD across participants (**Fig. S12**). Analysis of LFP patterns revealed that performance was contingent on the relative laterality in the encoding of muscle activation during walking.

### STN LFP encodes the vigor of leg muscle synergies

We next asked whether the encoding of vigor is a general principle that also applies to the activation of leg muscle synergies during walking. We translated the single-joint paradigm wherein participants were instructed to produce two levels of muscle activation to the context of walking. We requested participants to produce a sudden increase in step length during walking (**Fig. 6A**). Lengthening the step during transitions from a normal to a long step involved a significant increase in the amplitude of muscle synergies associated with ipsilateral propulsion and contralateral weight acceptance (Wilcoxon signed rank test, n=12, p=0.008).

**Figure 6.**
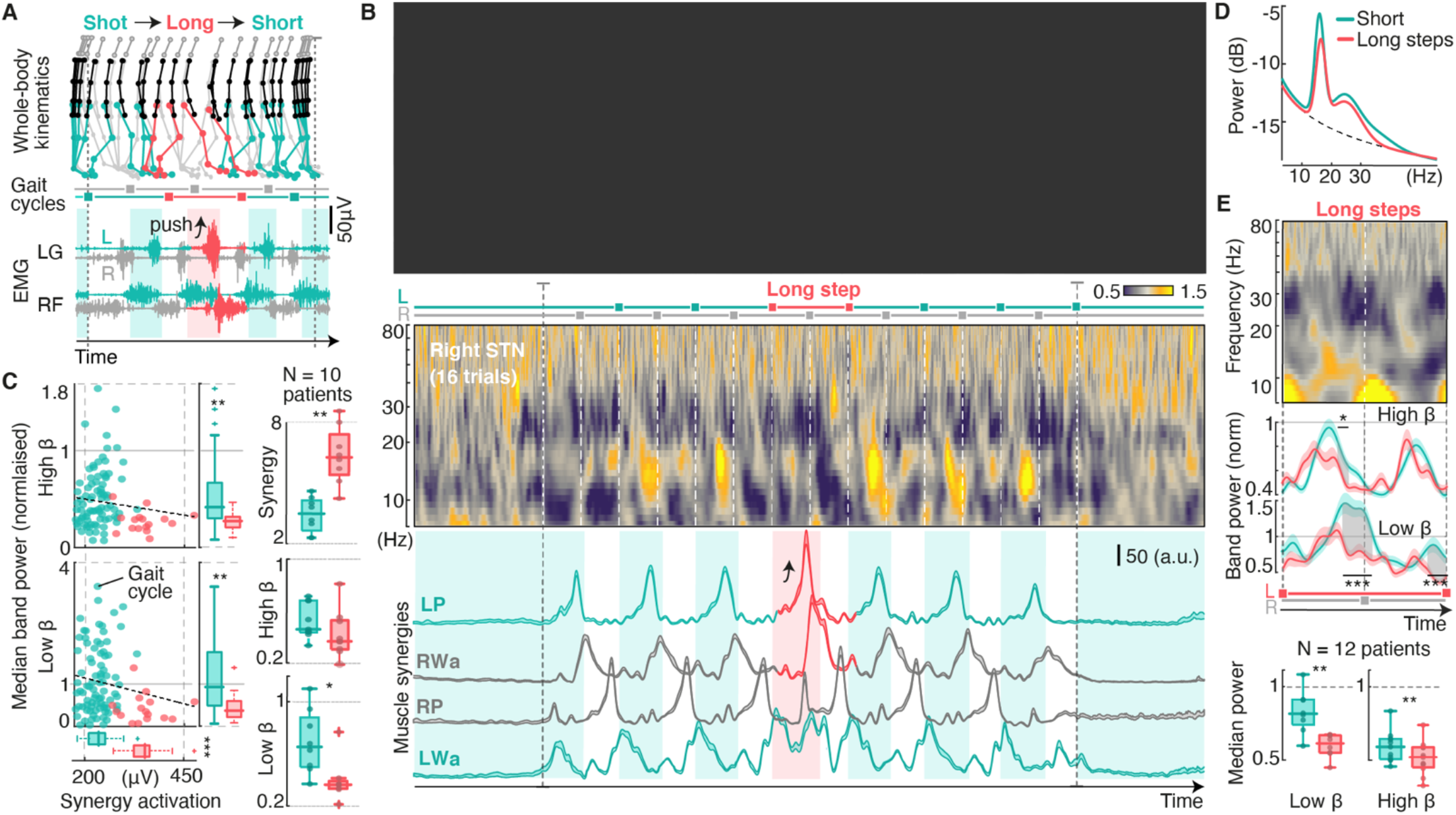
STN encodes modulations in force during walking. **(A)** Patients were instructed to perform a locomotor task that involved a long step interleaved during short steps. This required a bigger activation of muscles related to propulsion (leg in stance) and weight acceptance (landing on the contralateral leg). **(B)** Time decomposition of a complete walking sequence (patient P1). Chronophotography (top, please contact the corresponding author to request access to the image), average spectrogram (middle) and synergy traces (median ±SEM) highlight modulations in low- and high-beta aligned in time with the recruitment of contralateral propulsion (LP) and ipsilateral weight acceptance (RWa) synergies during the big step. **(C)** Correlation between band power (low- and high-beta) and synergy amplitude (sum of LP and RWa) for each gait-cycle. Steps requiring bigger synergy recruitment consistently exhibit strongly desynchronized low- and high-beta correlates (Wilcoxon signed rank test, * p<0.05, * * p<0.01). Boxplots showing cross-patient quantifications confirm this behavior. **(D)** Power spectral density during tasks requiring walking with sustained big steps, as compared to sustained small steps. Both low- and high-beta bands exhibit a reduction in power. **(E)** Time analyses per gait-cycle for this task. Average spectrogram (top) and power traces (median ±SEM, middle) aligned and interpolated between contralateral footstrikes. Predominant force-related changes occur just before footstrikes, aligned with propulsion phases. Boxplots showing quantifications across patients confirm this behavior (bottom) (Wilcoxon signed rank test test).

During the increase in step length, all participants exhibited a decrease in the power of both low- (signed rank test, n=8, p=0.023) and high-beta bands (n=9, p=0.055) (**Fig 6B**). This response abolished the rebound that occurred during late swing in low-beta and reinforced the desynchronization in high-beta power (**Fig 6C**).

We observed the same modulation patterns during walking involving sustained short versus long steps (**Fig 6D,E,F**).

### Continuous prediction of muscle synergy amplitude during walking

We then tested the possibility to predict the continuous modulation of leg muscle synergies from STN LFP, both during short and long steps. We established a deep learning model27 that takes as input the spectral decomposition of bilateral LFP signals and generates a continuous prediction of muscle synergy amplitude over time. The model was trained and tested on all recordings of walking with short and long steps (**Fig. 7A**). Since the power of STN LFP correlated with bilateral muscle synergies, we trained the model to predict the sum of all four muscle synergies. The model accurately predicted the amplitude (average R2=0.51) and timing (average cross-correlation = 0.30) of synergy profiles for all participants (**Fig. 7B,C,D** and **Fig. S14**).

**Figure 7.**
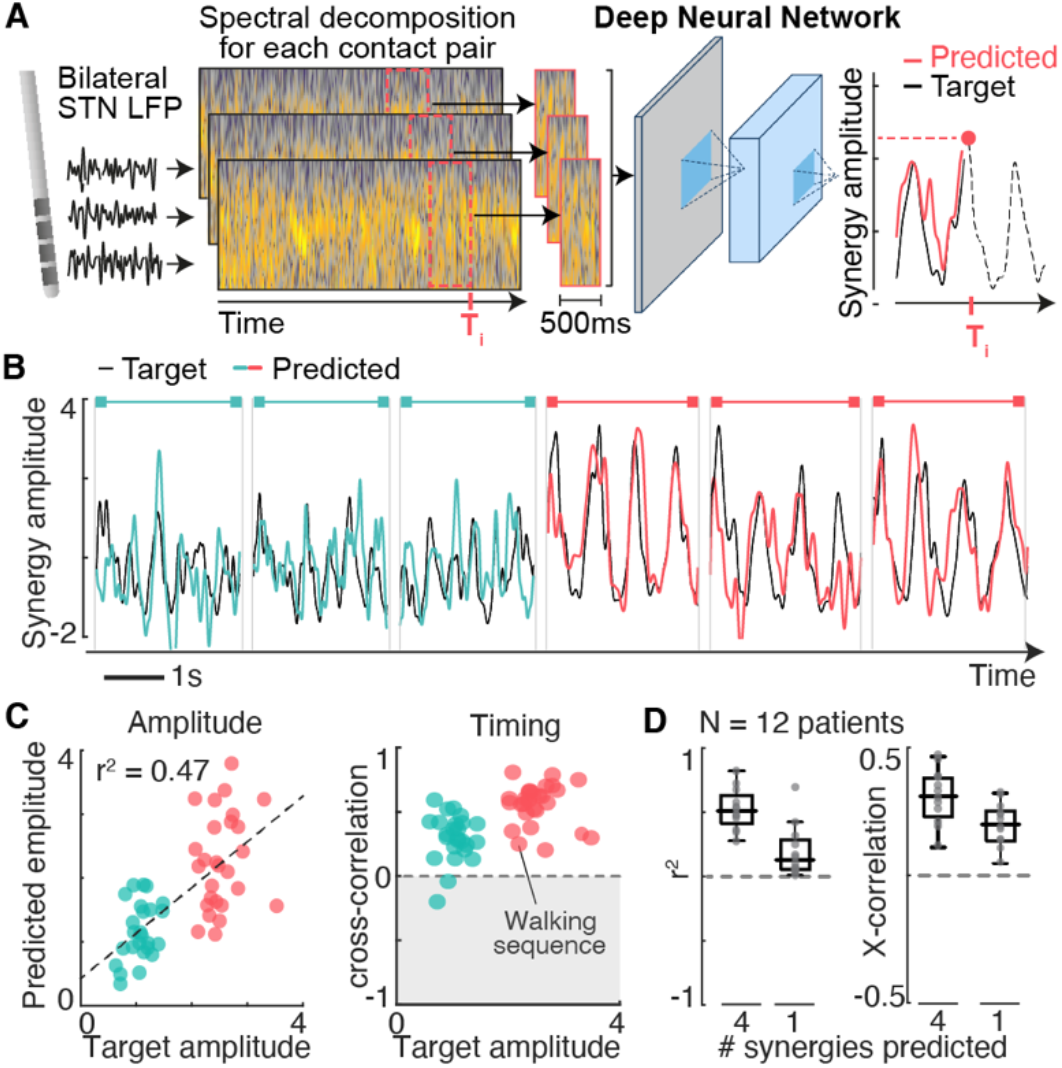
Prediction of muscle synergy activation during walking using a deep learning model. We implemented state-of-the-art deep neural network algorithms to model the continuous amplitude modulations of leg muscle synergies from STN LFP. **(A)** Architecture of the neural network. Bilateral raw LFP are pre-processed and used to compute a spectrogram for each contact pair (multitaper). For each timepoint (10ms resolution), a sliding window (500ms) is isolated and fed into a convolutional neural network (CNN) that processes their image properties. The CNN is composed of three consecutive convolutional layers with increasing receptive fields. Each layer identifies the spectrotemporal features that best predict the output at their specific resolution and pass them on to the next layer. A final layer (fully connected neural network) combines all features into a single prediction of synergy activation. Training and testing of the deep neural network are performed using leave-one-out cross-validation on all short and long walking sequences combined. We compared the performance when predicting an individual synergy (unilateral) versus predicting the sum of all four synergies (bilateral). **(B)** Illustrative examples of target vs predicted synergy traces for six walking sequences (three sequences of small steps and three sequences of big steps). **(C)** Quantification of performance when predicting the amplitude and timing (cross-correlation between target and prediction) of synergy profiles for each walking sequence. **(D)** Boxplot showing the cross-participant accuracy when modeling the sum of all (four, bilateral) synergy profiles vs predicting a single synergy.

Training the model to predict only one synergy led to lower accuracy, both in amplitude (average R2=0.21) and timing (average cross-correlation=0.24), compared to the prediction of all four synergies combined (**Fig. 7D**).

### Idiosyncratic encoding of walking

We found that participants exhibited both common and idiosyncratic features during single muscle activation. Importantly, comparisons of LFP patterns during walking versus single-joint paradigms revealed that these features were highly consistent across tasks for all participants (**Fig. S5 and S6**).

### STN LFP encodes muscle activation underlying activities of daily living

Activities of daily living require gait adaptations that involve modulations in muscle activation to accommodate leg movements to changing terrains. Therefore, we asked whether the encoding of muscle activation in the STN also captured these modulations.

We asked participants to walk naturally without any signs on the floor, and to step over an obstacle located in the middle of the room (**Fig. S15A** and **Movie S2**). Passing the obstacle required an increase in flexion to raise the foot above the obstacle, followed by an increase in propulsion contralaterally. These leg adaptations were paralleled by modulations in STN LFP that mirrored the patterns observed during the sudden production of a long step (**Fig. S15B)**.

While the decoder was trained on short versus long steps, the same decoder accurately predicted standing and walking without any visual cues, as well as modulations related to the obstacle (**Fig. S15C**).

### Decoding of freezing of gait

Finally, we sought to leverage our decoding framework to test whether dysfunctional components of walking could be predicted from STN recordings.

Four participants exhibited frequent episodes of freezing of gait, predominantly whilst turning. Two participants showed “complete akinesia”^28^ (E2 and P3), while the other two participants exhibited “leg trembling with little effective forward motion” (E3) or “shuffling steps with minimal forward movement” (P1). We asked a neurologist expert in movement disorders (A.Z.) to define these periods based on video recordings. To capture the precise time windows during which both feet were “glued to the floor”^28^, we combined these visually-defined periods with analyses of accelerometer data from sensors attached to the shoes of the participants (**Fig. 8A**).

**Figure 8.**
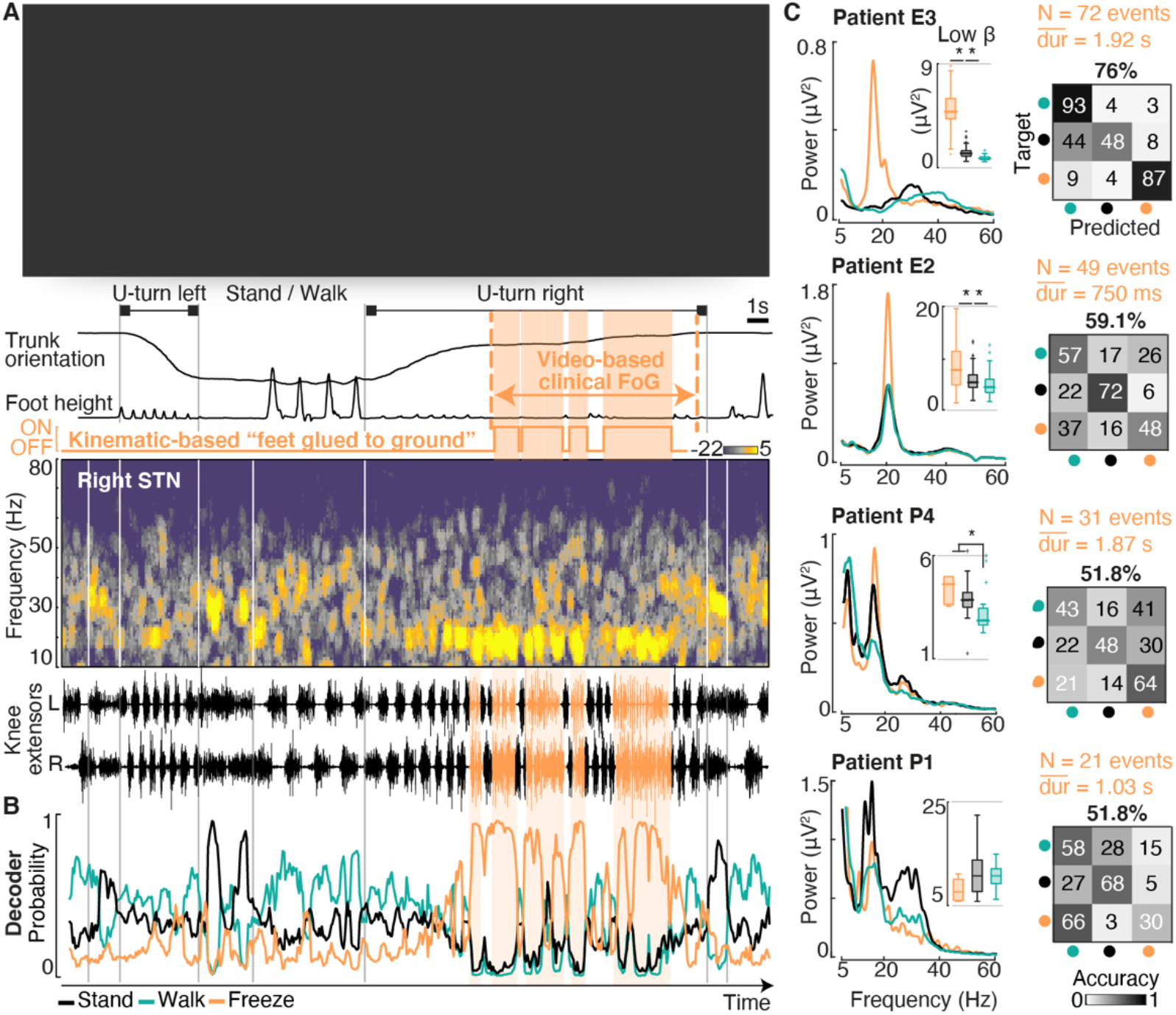
Prediction of freezing of gait episodes from STN LFP. **(A)** Illustrative example of a freezing of gait episode during turning. Chronophotography and kinematic traces of trunk orientation and foot elevation (top, please contact the corresponding author to request access to the image), spectrogram of raw STN LFP (middle) and bilateral knee extensor EMG traces (bottom) for a walking bout involving two U-turns. Periods of freezing of gait (defined based on video analysis by a movement disorders clinician) are indicated with orange dashed lines. Within those periods, we additionally computed the precise times during which feet are “glued to the ground” using kinematic sensors attached to the feet. **(B)** Aligned probability traces from a decoding algorithm trained to discriminate 3 classes (“stand”, “walk” and “freeze”). **(C)** Four patients exhibited episodes of freezing, which differed in their types28 and severity (duration and frequency of events). PSD profiles for each class highlight a predominance of low-beta power during freezing episodes that reflect the severity of FoG in each patient. Boxplots quantify the median power in the low-beta band for each class. We then trained our decoding algorithms to predict freezing episodes for each patient individually. Confusion matrices highlight the decoding performance for each participant.

Visual inspection of LFP spectral features revealed that these periods correlated with the emergence of predominant low-beta signatures in bilateral STN LFP, along with partial co-activation of knee extensor muscles that reflected the incapacity to shift bodyweight (**Fig. 8A**). Alternations of right and left muscle activation patterns resumed as soon as low-beta synchronization vanished.

Analysis of spectral components showed increased low-beta power during freezing periods compared to walking or standing, although the predominance of these signatures mirrored the severity of freezing events in each participant, and the frequency and duration of events^28^ (**Fig. 8C**).

We finally configured our machine learning classification platform to predict the occurrence of periods during which feet are “glued to the ground” (**Fig. 8B,C**). Cross-validation performance reached accuracies (between 60 and 75%) for participants exhibiting a large number of freezing episodes. Performance was variable for participants with few episodes to train the decoder (around 50% across three classes).

## DISCUSSION

We conceived a neurorobotic platform that allowed us to uncover fundamental principles of leg movement encoding in the STN. We found that the STN encodes the initiation, termination and vigor of leg muscle activation, independently of the recruited muscle. We found that the same principles underlie the encoding of bilateral leg muscle synergies during walking. We translated this understanding into machine learning classification algorithms that predicted walking states, gait events, vigor of walking, and freezing episodes. Here, we discuss the similarities in the principles that determine the encoding of arm and leg movements, and how understanding patient-specific modulations during isolated leg movements allowed the interpretation of functional and dysfunctional LFP patterns during walking.

### Encoding of sensory feedback in STN LFP

Sensory feedback is an essential source of control during walking29. People with PD exhibit a reduction in gait automaticity30. Consequently, studies suggested that the cerebral cortex may be more engaged in the control of walking in people with PD compared to healthy individuals31. Difficulties to integrate proprioceptive information may be responsible for this reduced automaticity32, to which basal ganglia dysfunction is likely to contribute. Experiments in non-human primate models and intra-operative recordings in humans showed that PD alters the encoding of sensory feedback in the STN33,34. We found that proprioceptive sensory feedback from the legs is also encoded in STN LFP. This encoding followed similar modulation profiles that emerged in the same frequency bands as those underlying volitional muscle activations. Together, these results suggest an involvement of the STN in the integration and processing of leg sensory feedback signals, especially proprioception.

### Encoding of gait initiation and termination in STN LFP

The basal ganglia has long been associated with the selection of motor programs2,8 and transitions between motor states7,10. The well-controlled conditions of our neuro-robotic platform allowed us to uncover systematic modulations in STN LFP during the initiation, termination and concatenation of leg muscle activation. We then translated these observations to the context of walking. We found that STN LFP encodes the initiation and termination of walking, as well as transitions between walking states when accommodating locomotor movements to sudden behavioral constraints, such as avoiding an obstacle. These results are compatible with observations in animal models. Single-unit recordings combined with activation/inactivation experiments showed that neurons located in various regions of the basal ganglia communicate with brainstem locomotor regions35,36 to control gait initiation11,37 and termination12,38.

### Encoding of bilateral leg muscle synergies in STN LFP

Previous studies reported that STN LFP encodes bilateral components of arm, hand or even finger movements3,39–41. This encoding did not appear to be specific for the type of movements or muscles involved. Our observations during isolated leg movements confirmed that the same principles apply to the encoding of leg muscle activation.

During walking, STN LFP displayed modulation patterns that directly reflected the activation of synergistic groups of leg muscles. These modulation patterns included two desynchronizations in beta power per gait-cycle that coincided with the activation of propulsion and weight-acceptance muscle synergies, followed by resynchronizations (and rebounds) during the ballistic component of the swing phase. These patterns emerged during steps from both legs. Deep learning modeling confirmed that the relative amplitude of bilateral muscle synergies is continuously encoded in the STN.

Our observations suggest that the STN encodes leg muscle activation during walking with high temporal resolution but low specificity. This conclusion is in line with current views about the role of the basal ganglia in the selection of motor programs that are encoded in other neural structures^42^. For example, it is well-established that the spinal cord encodes leg muscle synergies^43,44^. In turn, the modular activation of muscle synergies simplifies the elaboration of various locomotor programs^45^. Our results are compatible with the idea that the STN contributes to selecting muscle synergies during walking.

### Encoding of vigor in STN LFP

Single-joint experiments revealed that STN LFP encodes the vigor of leg muscle activation. However, this principle could only be detected during the sustained activation of muscles with different levels of force. Maintaining a strong force required continuous adjustments in muscle activation that were not required when maintaining a weak force. We surmise that these continuous adjustments imposed a continuous engagement of the STN to send corrective commands downstream, which delayed the resynchronization in beta power. The same principle applied during walking. The execution of long steps involved an increase in the amplitude of the muscle synergy associated with propulsion, compared to short steps. This increase abolished the rebound in low-beta that was observed during the ballistic phase of swing when participants walked with short steps.

Patients with PD display small shuffling steps and slow walking with reduced propulsion. While many aspects may contribute to these deficits, the difficulty to maintain a sustained desynchronization during the activation of propulsion-related muscles may partly explain these deficits. Consistent with our observations, experiments in animal models showed that the relative activity of basal ganglia neurons scale up with the speed of walking2,8,35,46. Similarly, the vigor and speed of movement was found to be encoded in STN LFP of patients performing manual tasks3,4,40.

### Automatic predictions of gait features from STN LFP

Understanding the principles through which leg muscle activation is encoded in the STN allowed us to configure machine learning algorithms that predicted gait features from STN LFP. We could predict walking states, gait events and vigor with accuracy. While each algorithm was trained and tested separately on individually defined classes, they could easily operate in parallel as part of an integrative platform for real-life applications.

However, we also identified limitations in the scope of gait features that may be predicted from STN LFP to control closedloop therapies. For example, spatiotemporal stimulation of the lumbar spinal cord^49^ has the potential to alleviate gait deficits in people with PD. This intervention requires real-time control of stimulation bursts targeting specific joint movements. Our results suggest that this spatial resolution is not accessible from STN LFP. Other brain regions may complement STN decoders for applications that require muscle-or joint-specificity. Increasing the resolution of STN recordings may resolve this issue. For instance, single-cell recordings reported preferential firing of STN neurons with different joystick directions in intra-operative conditions^47,^ as well as with detailed kinematics in animal models^48^.

### Clarification of conflicts in the literature

The encoding of gait in the STN has remained controversial. A subset of studies reported a reduction either in high-beta power^18,19^ or in the entire beta band^25,49^ during walking compared to standing, as well as frequency shifts in the beta peak^20^. Instead, other studies failed to detect changes in power^21^. Additionally, two studies documented beta modulations aligned to the phases of gait^18,19^, but the temporal patterns of these analyses did not match.

To disentangle these heterogeneous results, we implemented three strategies. First, we used an unbiased fitting algorithm to identify patient-specific frequency bands. Second, we informed the design of locomotor tasks based on the deconstruction of gait features during single-joint paradigms. Third, we computed explicit metrics of leg muscle activation that allowed us to establish direct links between the modulation of STN LFP and motor performance. These strategies uncovered systematic and reproducible correlations between STN modulations and the timing and vigor of leg muscle synergies. These strategies were essential to expose these principles. Instead, the considerable variability across patients hindered most of these correlations when LFP signals were averaged across gait cycles and patients. We infer that these idiosyncratic modulation patterns are largely responsible for the variability observed across previous studies.

### Encoding of gait freezing

Freezing of gait remains an enigmatic phenomenon. The ability to predict the occurrence of freezing episodes is essential to design therapies able to prevent or alleviate these deficits. Studies that investigated this phenomenon reported a variety of correlates in STN LFP, including increases in entropy^49^, low-beta power^25,26^ or cortico-STN decoupling^50^. Our recordings confirmed that gait freezing was associated with an increase in low-beta power, and that this increase coincided with periods when both feet were “glued to the ground” whilst turning. Concomitantly, abnormal activation patterns emerged in bilateral knee extensor muscles, which prevented patients from shifting body weight from one leg to the other despite the intention to turn.

We leveraged this understanding to design classification algorithms that predicted gait freezing, suggesting that our computational framework could discriminate between functional and dysfunctional walking states. However, the robustness of the decoders was variable across patients. Concretely, we found that the magnitude of low-beta modulations mirrored the type and number of freezing events. So did decoding performance. For patients who suffered infrequent episodes of gait freezing that terminated rapidly, we could not collect a sufficient amount of data to train and test the decoder. However, we anticipate that monitoring STN LFP throughout the day will enable the collection of the necessary data to train robust decoders.

We exposed fundamental principles through which the STN encodes leg muscle activation during isolated leg movements and walking. We predict that this understanding will open new possibilities to design algorithms that decode gait deficits in people with Parkinson’s disease. These predictions will support strategies that translate continuous recordings of STN LFP into control commands to operate neuromodulation therapies in closed loop.

## MATERIALS AND METHODS

### Study design

The objective of this study was to uncover the STN correlates underlying locomotor functions, and to leverage this understanding to develop decoders that predict functional and dysfunctional gait.

20 patients with Parkinson’s disease were recruited for the study. All participants received bilateral deep brain stimulation (DBS) leads (Medtronic 3389) and were recorded in the five days that followed their surgery. One participant, who was only able to perform single-joint experiments during the post-surgery period, was additionally recorded 6 months later on the walking tasks. Ten patients were recorded while their leads were externalized (referred to in the text as “patient E^x^”). Ten patients were implanted with a Percept PC stimulator (Medtronic, USA) in the right abdominal area, and were recorded using the sensing capabilities offered by the device (referred to as “patient P^x^”). Patients were predominantly in the OFF-medication condition (>12h), although depending on their severity, some retained some dopamine agonist medication (see **Table S1)**.

Not all patients participated in all motor tasks. 19 patients performed the isometric single-joint force tasks in the neurorobotic platform. Five of them additionally performed the isotonic and passive experiments. 15 patients performed the walking tasks (small, big and mixed steps). Four patients additionally performed the obstacle-avoiding task.

For single-joint experiments, two patients were excluded due to technical problems during data acquisition. For gait experiments, one patient had to be removed due to difficulties in synchronizing neural and kinematic data. Another patient was excluded because all LFP channels exhibited artifacts (see artifact definition below).

Overall, 17 patients were retained for analyses on the neurorobotic platform and 12 for gait experiments.

All experiments were approved by the Ethical committee of the Canton de Vaud, Switzerland. Informed consent was obtained after the nature and possible consequences of the studies were explained.

### DBS electrode localization

Lead localization was computed using the processing pipeline (Horn &Li et al. 2018) in the Lead-DBS Matlab toolbox (Horn &Kühn. 2017), using pre-operative T1 and T2-weighted MRIs and a post-operative CT-scan as inputs. Briefly, post-operative CT scan was linearly co-registered to pre-operative MRI using advanced normalization tools (ANTs, Avants et al. 2011). Co-registrations were visually verified and manually corrected if needed. A brain shift correction step was applied, as implemented in Lead-DBS. All preoperative volumes were used to estimate a precise multi-spectral normalization to ICBM 2009b NLIN asymmetric space (Fonov et al. 2009) applying the ANTs SyN Diffeomorphic Mapping (Avants et al. 2008) using the preset “effective: low variance default + subcortical refinement”. DBS contacts were automatically pre-reconstructed using the phantom-validated and fully automated PaCER method (Husch et al. 2017). They were all individually verified.

### LFP recordings

#### Externalized patients

Local field potentials were recorded by connecting the externalized extension leads to a high-resolution amplifier (Tucker Davis Technologies, USA for patients E1-3, and ANT-neuro, Netherlands, for patients E4-10). Digitized signals were imported into a dedicated computer. For patients E4-10, we used the OpenVibe library (Renard et al., 2010) to enable real-time acquisition of LFP. For each hemisphere, signals from each contact were referenced to the upper-most electrode. They were later re-referenced as bipolar channels between neighboring contacts. Recordings were high-pass filtered (2^nd^-order Butterworth filter, 3Hz cut-off), 50-Hz notched, and downsampled to 1kHz. Signals were visually inspected and whenever needed, artifacts were identified and replaced by a noise signal carrying the same frequency components as the whole recording. Synchronization with additional devices was performed using the “trigger” input to the amplifier.

#### Percept PC

Local field potentials were recorded using the sensing capabilities of the Percept PC (sampling frequency 250 Hz). Recordings in resting conditions (sitting or standing) were performed in the *Indefinite Streaming* mode, which records all 3 bipolar contact pairs. Recordings during motor tasks were obtained in the *Brainsense Streaming* mode, which is restricted to one contact pair per hemisphere. Prior to the first recording session with each patient, we performed a *Brainsense Survey* and visually inspected the PSD of each pair. We selected the contact pair with the highest beta power and kept that pair for all subsequent experiments. We further ensured that the selected contact pair was not labeled as “artefacted” by the system. Synchronization with external devices was performed by applying a transient DBS burst (130Hz, 60us, 1mA) at the beginning and end of each recording, which induced artifacts in an external electromyographic sensor placed on the chest of the patient, in the vicinity of the IPG (Thenaisie et al., 2021).

All recordings were performed in the OFF-stimulation condition. For Percept PC recordings in the *Brainsense Survey* mode, stimulation was ON at 0mA.

### Biomechanical recordings during gait

#### Kinematics

Externalized patients were recorded using a wholebody suit endowed with 19 motion sensors on key landmark joints of the legs, arms, trunk and head (Rokoko, Denmark). The suit transmitted accelerometer and magnetometer data wirelessly (100Hz) to a dedicated computer running the *Rokoko Studio* software, which reconstructed 3D body positions using an inverse-kinematics model. The model was personalized for each patient (definition of body height and segment lengths) on the first day of experiments. Prior to every recording, the initial pose of the model was calibrated during a baseline “standing” position to maximize the accuracy of subsequent movements. Patients implanted with the Percept PC were recorded in a gait lab using an optoelectronic motion capture system (Vicon, UK) that measured 3D positions of key body joints.

Kinematic data was complemented with bilateral triaxial inertial measurement unit (IMU) sensors (Delsys, MA, USA) attached to the patients’ shoes, which recorded raw gyroscope signals from the right and left feet (sampling frequency: 148Hz). All recordings were synchronized using dedicated trigger signals.

#### Electromyographic signals

EMG signals were recorded using a wireless system operating at 1.5kHz (Noraxon, USA). Sensors were placed bilaterally, on agonist and antagonist muscles of the ankle joint (TA Tibialis Anterior, MG Medial Gastrocnemius, LG Lateral Gastrocnemius) knee joint (VM Vastus Medialis, ST Semitendinosus) and hip joint (RF Rectus Femoris). For patients implanted with a Percept PC stimulator, an additional EMG sensor was placed on the chest for synchronization purposes.

### Identification of LFP power frequency bands

The identification of power frequency bands was personalized for each patient. Theta, low- and high-beta, and gamma bands were identified using an unbiased algorithm that fits the PSD of each patient as a combination of an aperiodic (1/f) and a mix of Gaussian components^24^. The algorithm iteratively optimized the fit and found the most appropriate number of Gaussian components and their parameters (mean and std) to model the original spectrum with a given accuracy.

Since frequency bands may differ or shift^20^ between rest and movement, and specific bands may only emerge during movement (such as gamma), we run the fitting algorithm on the PSD computed on the complete recordings in the neurorobotic platform (isometric task), which included both resting and leg effort periods. Identified Gaussian components in the 13-20 Hz range were labeled as “low-beta”, and those in the 20-35 Hz range were labeled as “high-beta”. All 17 participants exhibited a high-beta band. 10 participants did exhibit a low-beta band (**Fig. S5 and S6**).

### Joint-specific motor tasks in the neurorobotic platform

Patients were comfortably sitting on an instrumented isokinetic machine (HUMAC NORM, CSMI, USA). The shin of their most affected leg was strapped to a pushing lever, connected to a dynamometer that measures rotation angles, speeds, and forces applied during a range of experimental conditions (isotonic or isometric exercises, active or passive movements). A screen provided patients with real-time visual feedback about their performance during each task.

#### Active isotonic movements

Patients were asked to perform a knee-extension movement from a resting position (baseline, 90-degree angle of the knee joint) to a full leg extension, and to release the leg back to baseline. 30 repetitions were recorded, interleaved with pauses of 5s duration.

#### Passive movements

Patients were asked to completely relax their leg (at the baseline position) and to not exert any resistance. The neurorobotic platform was programmed to passively move the patients’ leg with an angular range and speed matching the ones recorded during active isotonic movements. Each trial was interleaved by 5s resting periods.

#### Isometric transient effort exercises

Patients were asked to push against the lever and to exert a predefined isometric force (either a low or a high force, defined as 33% and 66% of their maximal voluntary contraction respectively). Throughout each trial, they received visual feedback of the force being exerted, along with the target. They were asked to release the effort immediately as soon as they reached the target (displayed on the screen as a yellow circle with predefined diameter, it disappeared from the screen as soon as patients reached it).

#### Isometric sustained effort exercises

Similar to the previous task, but patients were asked to reach and maintain a low or a high force for five seconds before releasing their effort. The target remained displayed on the screen for the duration of the task and patients were instructed to maintain their force within the circle.

### Locomotor tasks

#### Walking tasks (small and big steps)

Patients were instructed to stand for approximately 3 seconds before initiating a sustained bout of walking at their comfortable speed (in a straight line), placing their feet on marked lines on the floor. Marked lines were spaced either 47cm (small) or 70cm (big) apart from each other. When arriving at the end of the bout, patients were instructed to stop and stand for another 3 seconds, before doing a U-turn and starting again.

#### Obstacle task

Patients were asked to walk at their natural speed in a straight line (without any marks on the floor). An obstacle (height 10.5cm, length = 39.5cm) was placed on their path. They were requested to stride over it and continue walking normally until the end of the bout.

### Identification of gait events &Definition of locomotor states

#### Bilateral gait events

We computed periods of walking and turning, along with toe-off and heel-strike events, automatically from gyroscope signals from the right and left feet using a modified version of the two-step framed threshold algorithm^24^. All identified events were validated manually. Briefly:

First, uninterrupted walking sequences were identified from low-pass filtered signals in the sagittal plane (3Hz, 5^th^-order Butterworth). Mid-swing peaks were detected using a threshold (defined as the average of the top ten peaks and scaled by 0.2. A minimum value of 40 degrees/seconds was taken) and pooled together when less than 3 seconds apart. Peaks of right and left sensors that did not occur consecutively were discarded. Identified walking sequences lasting less than 5 seconds and occurring during concomitant peaks in the coronal plane (which corresponded to turning periods, see below) were also removed.

Within each walking sequence, we extracted heel strike and toe off events. Gyroscope signals were low-pass filtered at 5Hz (5^th^-order Butterworth). We first refined the identification of mid-swing points. These points needed to be at least 0.5 seconds apart, and greater than a sequence-specific threshold (defined as the mean of all points exceeding the sequence mean). Any mid-swing points not complying with these rules were discarded. We then computed “full contact” times, i.e. when feet were in full contact with the floor (these corresponded to a plateau in the acceleration profiles). They were identified as the maximum of the plateau between consecutive mid-swing peaks. From there, (i) heel strike events were identified as the minima occurring between mid-swing and “full contact” points, and (ii) toe-off events were defined as the minima occurring between “full contact” points and the next mid-swing points.

We then identified turning sequences. The low-pass filtered gyroscope signals in the coronal plane (5Hz, 5^th^-order Butterworth filter). Turning peaks needed to be superior to a threshold (defined as 75% quantile of the datapoints exceeding the mean of the whole recording) and at least 0.2 seconds apart. Turning sequences were pooled together: Peaks separated by less than 1.75x the mean interpeak distance throughout the recording were assumed to belong to the same turning sequence.

#### Locomotor states

Using the aforementioned gait events, we categorized each time-point of the recordings into five discrete locomotor states, labeled as “standing”, gait “initiation” and “termination”, “continuous walking”, and “turning”.

For each walking sequence, gait initiation was defined as starting 0.5s prior to the first heel-off (time needed for the postural adjustment and bodyweight shift required to lift the leg) until the first heel strike. Gait termination was defined as the last 2 gait cycles (one full step with one leg, and a last one to bring the trailing leg next to the leading leg). Continuous walking corresponded to all steps in between. Standing was defined as the periods between the end of turning and the beginning of gait initiation (standing pre-walk), or as the period between gait termination and the beginning of turning (standing post-walk).

This state-machine description of gait was further used for developing gait-state decoders and compute state-related analyses of power.

#### Computation of muscle synergies

We first computed EMG envelopes for each individual muscle. Raw EMG signals were band-pass filtered (20-500Hz, zero-lag 4^th^-order Butterworth filter), full-wave rectified and smoothed (zero-lag 4^th^-order low-pass Butterworth filter at 7 Hz). Envelopes were normalized so that their maximum would be one.

Synergies were derived from all muscle envelopes of the left and right leg together (N = 12 muscles), using recordings from both the small and big stepping tasks. A non-negative matrix factorization (NNMF) algorithm was iteratively run (1000 iterations beginning from 10 initial seed values, randomly selected using the multiplicative update algorithm). For each patient, the 4 first dimensions were kept, and we verified their percentage of variance explained (∼90% across patients). We then labeled each one of the extracted synergies as right or left “weight acceptance” or “propulsion” based on the muscle weights and their temporal activations profile.

To obtain average synergy activation profiles for each task, synergy traces were linearly time interpolated over the 4 phases of each individual gait cycle (bilateral stance, swing and double stance, as defined by gait events). Gait initiation and termination steps were excluded from this average. The area under the curve of for synergy was computed using trapezoidal numerical integration and compared between tasks across subjects.

The synergy sub-space identified on small and big steps was then also used to extract the temporal activation profiles of synergies for other tasks (e.g., mixed steps) by multiplying muscle envelope signals with the identified weights.

### Identification of artefacted LFP channels during gait

Gait-related artifacts affect signals predominantly in the low-frequencies but can also spread to higher frequencies, which makes them difficult to be removed through standard filtering. Cleaning methods using ICA or advanced signal processing tools have been reported in literature (Gwin et al., 2019) but results and interpretations have remained controversial (Castermans et al., 2014). Moreover, the identification of corrupted channels itself can be tricky, as movement-related neural modulations and artifacts are both locked to the rhythm of gait.

Rather than aiming to identify artifacts in the time-domain, we reasoned that corrupted channels would exhibit important differrences in the aperiodic (1/f) component of the power densities (PSD) between rest and walking: The aperiodic component captures the overall baseline power across the spectrum and should not change importantly over consecutive trials (task-related modulations are expected to be captured in the activity of periodic frequency bands)

For each patient, we thus applied the fitting algorithm^24^ to the PSD of two separate recordings, one at rest (sitting) and one during walking (from the same session). We then compared their aperiodic (1/f) component: We computed the root mean square error (RMSE) in the range [10-90] Hz (region of interest) and ensured that walking did not induce an increase in 1/f power bigger than 50% compared to that sitting (difference of 1.76 dB) (**Fig. S10**). All channels that exceeded that value were considered as corrupted in the region of interest and discarded for further analyses or decoding purposes. Visual inspection of the spectrogram for channels labeled as “artefacted” consistently showed important low-frequency spikes that periodically corrupted the spectrogram and spread to higher frequencies. All retained channels were also verified by visual inspection of their spectrogram.

We note that this approach is highly restrictive, in that channels may still convey useful information (for instance in high frequencies that have *a priori* not been corrupted) but are nonetheless completely discarded. Overall, N=6 participants exhibited at least one corrupted channel. One patient (implanted with the Percept PC) had to be completely excluded as all contacts (one per hemisphere) were artefacted.

### Spectral and power analyses of experiments in the neurorobotic platform

#### Identification of events and interpolation

Events were defined from knee angle (isokinetic tasks) or torque (isometric tasks). Start, hold, release and end events corresponded to the maximum (start) and minimum (stop) torque acceleration, and on angle/torque thresholds manually defined for each recording (hold and end). The baseline was defined as the 2.5s preceding *start*, and the post-effort resting period as the 2.5s following *end*.

#### Interpolation

Data (torque, leg angle, EMG envelopes, band power data and power spectrum) was epoched and interpolated for each time window defined by two events. Epochs containing bad task execution or LFP artifacts were manually excluded.

#### Average scalograms

Scalograms were computed with continuous wavelet transform (*cwt* function, Morlet’s wavelets) from 8 to 95Hz and log-normalized to baseline for each frequency as:

#### Band power

The whole recording was filtered to the band limits (Butterworth order 4), and power was computer after Hilbert transform. Powers were epoched and interpolated. Power of each epoch were log-scaled and normalized to the pre-movement resting period, and smoothed with a 0.5s moving average before mean and standard error of the mean computation.

#### Statistical analysis

Scalograms and band power were tested at the subject-level for difference between conditions (e.g. low and high forces) with Monte-Carlo cluster randomization (200 randomizations, maximum sum of t-scores). Band powers were further tested at the group-level by averaging across each time window for each patient. Differences to baseline were tested with t-test. Differences between conditions (e.g. low versus high force) were tested with non-parametric Wilcoxon test. Bonferroni correction was applied for the number of bands tested (n=2).

### Spectral and power analyses of gait experiments

#### Raw spectrograms

Spectrograms of raw LFP signals were computed through multi-taper decomposition using the Chronux library (http://chronux.org/) between 5 and 125Hz (N = 3 tapers, 1s window with 50ms overlap).

#### Average scalograms

Average scalograms were computed both for complete walking sequences (starting 2s prior to the first heel off and lasting until 2s after the last footstrike) or per gait cycle (between consecutive footstrikes of the same leg). For both cases, the scalogram of all the recorded sessions for each condition were computed using continuous wavelet decomposition (Morlet wavelets, Matlab, USA). Each frequency was normalized with respect to the mean power computed during “standing” periods (as previously defined). Walking sequences were pooled together depending on whether they started with the right or left foot, interpolated (5000 points for the standing periods before and after walking, and 1500 points between consecutive footstrike events during stepping) and averaged over time. For gait-cycle averages, consecutive steps were pooled together after interpolation and averaged in time.

#### Computations of band-power per gait states

For each frequency band, power was computed from the bandpass filtered LFP signals ((4^th^-order Butterworth filter, limits defined by the fitting algorithm). Power was normalized with respect to the mean power of all “standing” periods pooled together.

To compare band-power changes per gait state (standing, walking, initiation, or termination), we computed the median power for each repetition of each state (N = 20 sequences) and performed nonparametric statistical analyses for each patient (Kruskal-Wallis test followed by multiple comparisons with Bonferroni correction).

#### Cross-correlation between band-power and synergies

For each gait-cycle (defined as the period between consecutive footstrikes of the same leg), we iteratively computed the crosscorrelation coefficient between the z-scored band-power and the z-scored synergy profile (using the sum of all 4 synergies).

#### Comparisons of power per gait-cycle

Comparisons in bandpower between short versus long steps were computed by comparing the time-interpolated power traces of each band (between consecutive footstrikes) using Monte-Carlo cluster analysis (200 randomizations, sum of t-scores). To derive statistics across-patients, we computed the median power of each band for each gait cycle. We restricted the calculations to a window of 200ms before each footstrike as key differences had been observed in those periods.

### Decoding algorithms of force production during single-joint experiments

#### Single-joint force decoder -- offline training and testing

For externalized patients, raw LFP signals (6 channels, 8kHz sampling frequency) were low-pass filtered at 1kHz and downsampled to 2 kHz. For patients implanted with the Percept PC, raw signals (2 channels, 250 Hz sampling frequency) were used without further pre-proce-ssing.

To train each class, we defined an epoch range of 2 seconds relative to the onset of each class (the “rest” class started 2s before the start of the movement, the “weak force” and “strong force” classes started when participants reached the target force and started holding a constant force). Within each epoch range, we iteratively estimated the PSDs of the LFP over sliding windows of 500ms with 30ms overlap. Each PSD was computed in a predefined frequency range (1-150 Hz for externalized patients, 1-125Hz for Percept PC) using a multi-taper method (2Hz per bin, Thomson 1982), as implemented in the Python MNE library (Gramfort et al., 2013). All PSD vectors from all channels were then concatenated into a unique feature vector of size = Nbins x Nchannels (75×6 = 450 dimensions for externalized patients, 60×2 = 120 dimensions for Percept PC).

We used a custom-built Python software for real-time decoding (https://github.com/dbdq/neurodecode). The decoder was based on Random Forest algorithms and implemented using the python Scikit-learn library (hyperparameters: 1000 trees, 5 maximum depth, and balanced subsampling). Testing performance was computed using Monte-Carlo cross-validation over 20 repetitions (20% of data for testing, 80% for training) within the epoch windows. Feature importance vectors (contribution of each frequency band to the overall prediction) were calculated for each channel from the trained model, and normalized (sum of all contributions equal to 1). Quantification of performance (**Fig. 3** and **Fig. S8**) is reported as (i) median probability traces (interpolated) for each class, separately for weak and strong force levels, (ii) sample-based confusion matrix across all weak and strong trials, and (iii) median probability values of each class across all weak and strong trials.

#### Single-joint force decoder – online real-time experiments

All three patients tested in real-time were externalized (Patients E1, E2 and E3). The decoder was previously trained and tested offline using data recorded on the previous day. During real-time experiments, raw LFP data was pulled from the amplifier at 2kHz (one sample every 0.5ms) and buffered (buffer size of 500ms). PSD computations and predictions were computed at approximately 30 Hz (one prediction every 33ms) to comply with computational resources. Computed probability traces for each class were then exponentially smoothed (alpha=0.1) to prevent abrupt noisy peaks or artifacts.

### Decoding algorithms during gait experiments

#### Decoder of gait states

Decoders were trained to discriminate between “stand”, “walk” and “start/stop” classes. We employed only LFP signals from contacts that were not corrupted by movement-related artifacts (see definition above). Additionally, we only considered frequencies above 10Hz to ensure that only physiological data is accounted for (range used for PSD computations = 10-125Hz, for all externalized and Percept PC patients). Epoch ranges were reduced to 1 second (relative to the onset of each class), to account for the shorter duration of some classes during walking. The onset times of each class were defined as follows: the “standing” class started 1s before the “initiation” state and after the “termination” state (see definition of states above), the “start/stop” class started at the beginning of the “initiation” state and at the beginning of the “termination” state, and the “walk” class was defined within windows of 600ms around each footstrike (right and left combined) during the “continuous walking” state.

Quantification of performance (**Fig. 5** and **Fig. S11**) is reported as (i) median +/-sem probability traces (interpolated within consecutive footstrikes) for each class, separately for short and long steps, (ii) sample-based confusion matrix across all short and long tasks, and (iii) median probability values of each class of the entire duration of each state across all short and long tasks.

To further evaluate changes in decoder performance induced by artifact removal, we compared the features contribution when decoders were trained using all contacts (as compared to only using non-artefacted contacts) (**Fig. S10)**. Removed channels systematically showed predominant contributions of very low frequencies. Their removal did not importantly change the frequencies contributing from the other channel when starting at 10Hz.

#### Decoder of gait events

Decoders were trained to discriminate between “stand”, “right footstrike” and “left footstrike” classes. Electrode contacts and training parameters were identical to those used in the decoder of gait states. The onset times of the “standing” class were kept unchanged. The “right footstrike” and “left footstrike” classes were defined within a window of 600ms around each footstrike event. Quantification of performance (**Fig. 5** and **Fig. S12**) is reported as (i) median +/-sem probability traces (interpolated within consecutive footstrikes) for each class, separately for short and long steps, (ii) sample-based confusion matrix across all short and long tasks (computed within a window of 100ms around each event), and (iii) median probability values of each class of the entire duration of each state across all short and long tasks (same window).

#### Decoder of gait freezing

Decoders were trained to discriminate between “stand”, “walk” and “freezing” classes. Electrode contacts and training parameters were identical to those used in the decoder of gait states. The onset times of the “standing” class and the “walk” class were kept unchanged. The onset times of the “freezing” class were defined based on the times of “feet glued to the ground” (see FoG definition below). Quantification of performance (**Fig. 8**) is reported as sample-based confusion matrix across all recorded trials for each patient.

### Deep neural network algorithm for the prediction of continuous levels of muscle activation

#### Architecture of the convolutional neural network (CNN)

Bilateral LFP were used to compute a spectrogram for each contact pair (multitaper). For externalized patients, raw signals were low-passed at 1kHz and downsampled from 8kHz to 2kHz. For Percept PC patients, signals were preprocessed with a hardware bandpass filter in the range of 0.5-100 Hz and sampled at 250Hz. At each timepoint (every 10ms), a spectrogram vector was estimated from a sliding window of 500ms. 100 consecutive spectrogram vectors were concatenated and fed into the CNN as an input sample with the mapping muscle activation value as the target output.

The CNN was composed of 3 consecutive one-dimensional temporal convolutional layers with increasing receptive fields (layer1: 34 filters with receptive field of view (FoV) of 30ms, layer2: 64 filters of 60ms FoV, layer 3: 128 filters of 120ms FoV). Each layer was trained to identify the spectro-temporal features that best predict the output at their specific resolution, which were passed to an average-pooling layer with a temporal stride of 2. The output values from the final pooling layer were then flattened and fed into a fully connected layer of 512 nodes, which are finally fed into a single output node without activation function that is mapped to the normalized muscle activation value. Training and testing of the deep neural network were performed using leave-one-trial-out cross-validation on all short and long walking sequences combined. Cross validation was performed at the trial level, i.e. samples were grouped by each trial so that samples from the same trial do not split into training and testing folds, to avoid having optimistic performance due to the temporal proximity of samples.

Performance (**Fig. 7** and **Fig. S14**) was quantified in terms of (i) the capacity to predict changes in synergy amplitude, quantified as the 95% quantile of each walking sequence, and (ii) the timing of synergy modulations, computed as the cross-correlation between the target and modeled traces of each walking sequence.

### Definition of time-periods of freezing of gait

#### Clinical evaluation

Periods of freezing of gait were identified based on video analysis by a neurologist expert in movement disorders (A.Z).

#### Kinematic detection of feet “glued to the ground”

Within the periods identified by the neurologist, we computed the times in which the feet of patients were “glued to the ground” using inertial sensors attached to the shoes of the participants. We applied a threshold (1.5x the mean) to the norm of the 7Hz low-pass filtered accelerometer data of each foot. This defined a binary vector (1 when a foot is moving, 0 when it’s static). We added right and left vectors and only preserved states equal to 0 (no movement from either feet) lasting more than 0.5s.

#### Computation of PSD and band-power during freezing episodes

For each state (stand, walk or freezing), we concatenated all occurrences and computed the overall PSD using Welch method (sliding window of 1s with 50% overlap).

### Statistics

Experimental data and simulations were processed offline using MATLAB R2018b (MathWorks). All data are reported as mean or median values, ±SD or SEM, as indicated. Normality of data was tested using a Kolmogorov-Smirnov test with 95% confidence interval (CI). Comparisons between conditions were computed using one-tailed t test (parametric) or Wilcoxon sign rank test (non-parametric) with 95% CI. Multi-group comparisons were performed using one-tailed ANOVA (parametric), or Kruskal-Wallis (non-parametric) tests with 95% CI, followed by Bonferroni correction.

## Data Availability

Anonymized data that support the findings of this study are available from the corresponding author upon reasonable request.

## SUPPLEMENTARY MATERIALS

**Fig. S1**. Anatomical reconstructions of deep brain stimulation lead placement across participants.

**Fig. S2**. STN LFP modulations during concatenation of force levels

**Fig. S3**. STN LFP modulations are irrespective of leg joint or movement direction

**Fig. S4**. STN LFP modulations are irrespective of ipsilateral or contralateral leg muscle activation

**Fig. S5**. Idiosyncratic encoding of leg muscle activation across tasks for participants implanted with a Percept PC stimulator

**Fig. S6**. Idiosyncratic encoding of leg muscle activation across tasks for participants recorded while their DBS leads externalized

**Fig. S7**. STN LFP modulations per gait-cycle

**Fig. S8**. Decoding performance of single-joint vigor tasks across participants

**Fig. S9**. Muscle synergies across participants

**Fig. S10**. Identification of artefacted channels during walking

**Fig. S11**. Decoder of walking states across participants

**Fig. S12**. Decoder of gait events across participants

**Fig. S13**. Performance of walking-state decoder during gait adaptations

**Fig. S14**. Model of leg muscle synergy profiles using a deep learning algorithm across participants

**Fig. S15**. STN LFP modulations during activities of daily living

**Table S1**. Clinical details for all patients

**Movie S1**. Neurorobotic platform to study the encoding of leg muscle activation in the STN

**Movie S2**. Encoding of leg muscle activation in the STN during walking

**Movie S3**. Decoding of walking features from STN local field potentials

## AUTHOR CONTRIBUTIONS

YT, KL, GC, JB and EMM conceived the study. YT, KL, CM, SS, AG, EP and EMM established the technological platforms and acquired the data. YT, CM and EMM performed electrophysiological and biomechanical data analyses. KL and SS built decoding algorithms and analyzed their performance. KL developed deep learning models and analyzed their performance. MB and JR designed 3D illustrations. EA, BW, AZ, MCJ, JFB and JB recruited patients and performed clinical evaluations. JB performed surgeries. YT, KL, CM, SS, MB, JR, JB, GC and EMM prepared the figures. YT, KL, GC, JB and EMM wrote the manuscript, and all authors contributed to its editing. AP, JB, GC and EMM acquired funding. EMM supervised all aspects of the work.

## ACKNOWLEDGEMENTS

We are grateful to J. Courtine for recording the voice-over for the videos, and to C. Varescon for help with figure editing and feedback on the manuscript.

## CODE AVAILABILITY

Custom code used in this study is available from the correspondding author upon reasonable request.

