## Supplementary materials for "Principles of gait encoding in the subthalamic nucleus of people with Parkinson’s disease"

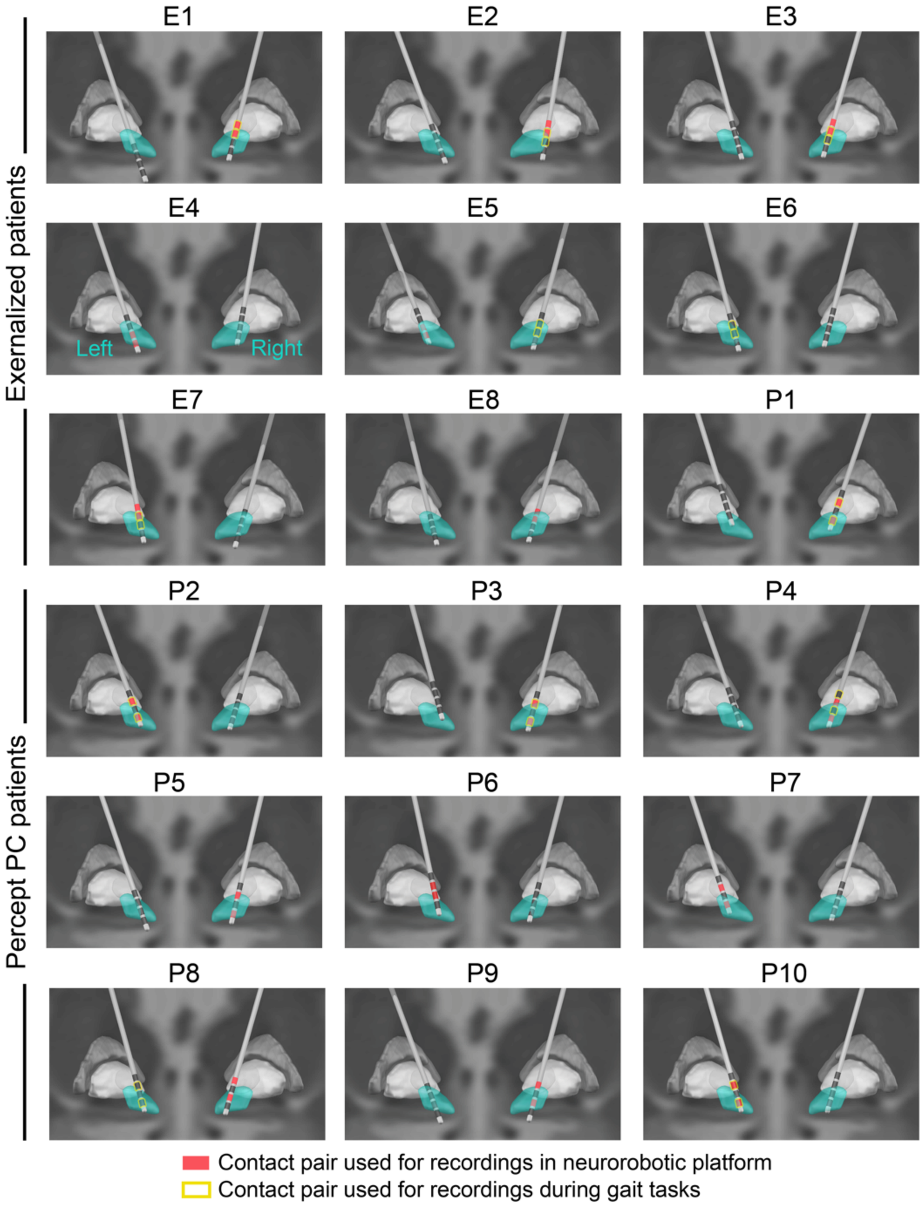


**Figure S1 I Anatomical reconstructions of deep brain stimulation lead placement across participants.** Recordings were systematically obtained from the most affected STN. Electrode pairs were kept identical for experiments in the neurorobotic platform and during gait, except for participants who exhibited strong gait-related artefacts in those channels: for externalised patients (three contacts per hemisphere), a nearby contact was chosen in such cases. For Percept PC patients (one contact per hemisphere), the other side was considered.

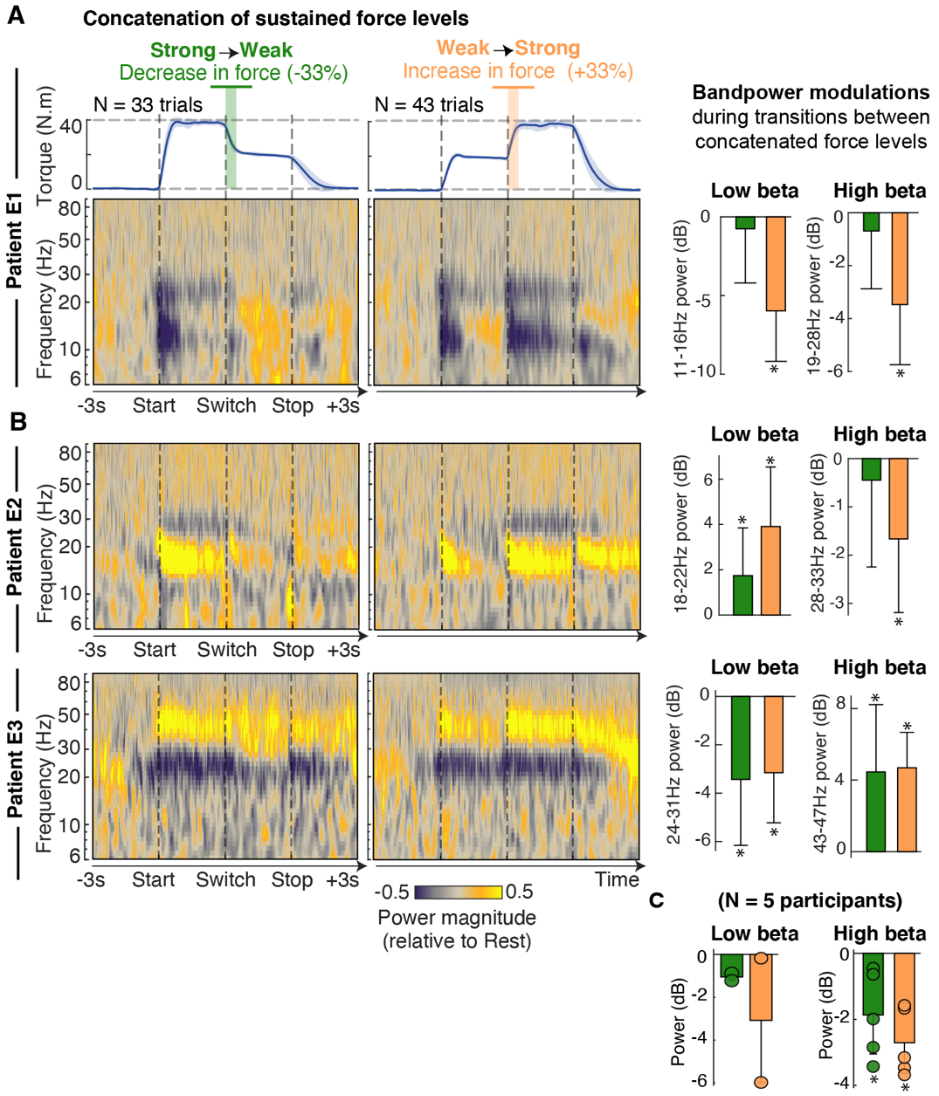


**Figure S2 I STN LFP modulations during concatenation of force levels. (A)** STN modulations during transitions between two different levels of sustained force, either from a weak to a strong force (left), or inversely (right). Average torque (mean ±SEM) and average spectrograms (rest-normalized, participant E1). Low- and high-beta de-synchronizations emerge at the initiation and the termination of muscle activation, as well as during the transitions between force levels. Barplots display band-power changes (mean ±SD) during transitions compared to baseline. Asterisks display significant differences with respect to baseline (t-test). **(B)** Similar representations for two other participants exhibiting different frequency band definitions and behaviors: Participant E2 exhibited an increase in 20 Hz power with movement, which scaled up with force. Participant E3 exhibited an increase in low-gamma power that modulated with force. **(C)** Low- and high-beta band-power (mean ±SD) across all participants who performed this task (N = 5). Despite clear patient-specific bands and behaviors, the encoding of transitions, states and vigor is present for all subjects in this well-controlled task.

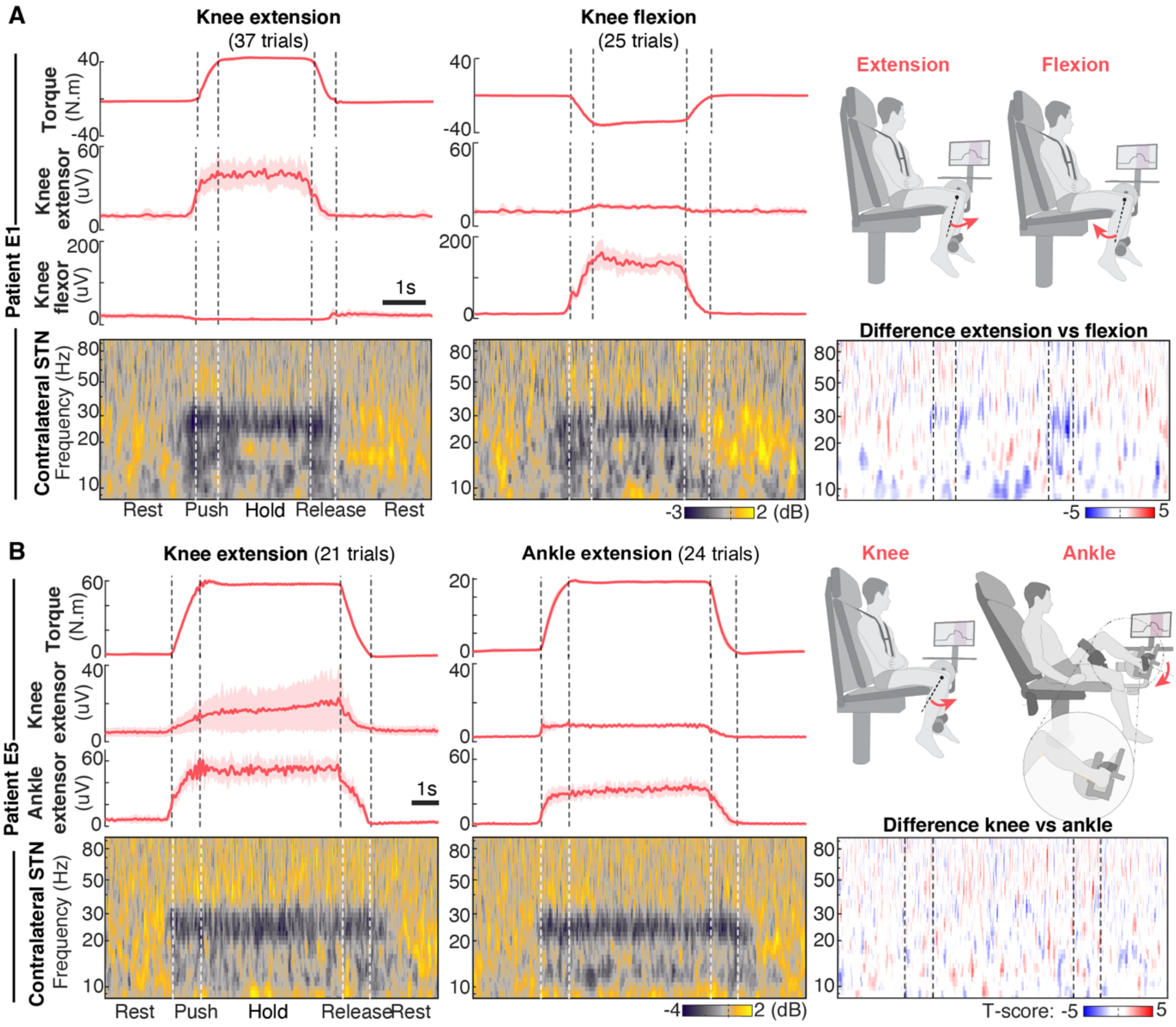


**Figure S3 I STN LFP modulations are irrespective of leg joint or movement direction. (A)** Patient E1 performed an isometric leg motor task restricted to the knee joint, either in extension or flexion, with two levels of force (low force 33% and high force 66% of maximal force in each direction). Profiles of torque (mean +/-SD), knee extensor muscle envelope (vastus lateralis) and flexor muscle envelope (semitendinosus), and scalograms of the contralateral STN (most affected hemisphere) normalized to pre-movement. Significance map (t-scores) of the difference between extension versus flexion scalograms (Monte-Carlo cluster randomization) identified no significant clusters in STN modulation despite the difference in movement direction. **(B)** Patient E5 performed an isometric leg motor task restricted to either the knee or the ankle joint, with two levels of force. Profiles of torque (mean +/-SD), knee extensor muscle envelope (medial gastrocnemius) or ankle extensor muscle envelope (rectus femoris RF) and scalograms left STN, normalized to pre- movement. Significance map (t-scores) of the difference between knee and ankle scalograms.

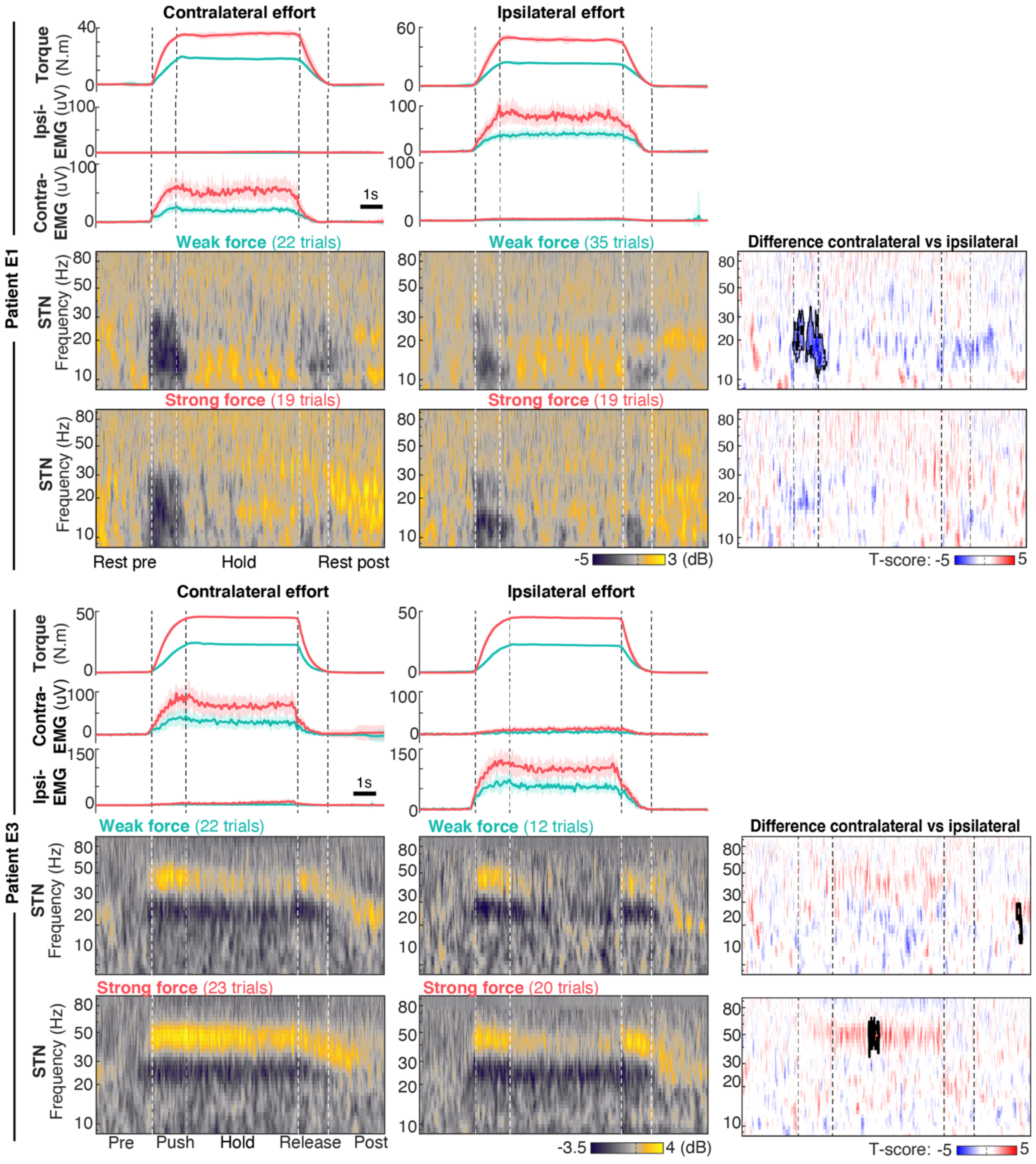


**Figure S4 I STN LFP modulations are irrespective of ipsilateral or contralateral leg muscle activation.** Two participants (E1, E3) performed an isometric knee extension task with each leg and with two levels of force. Weak and strong force levels were defined as 33% and 66% of the maximal voluntary contraction of each leg. Temporal profiles of torque (mean +/-SD), knee extensor muscle envelopes and scalograms of the most affected STN. Map of statistical difference (t-map) between contralateral versus ipsilateral movements revealed statistically different clusters, either in beta desynchronization (participant E1) or in low-gamma synchronization (participant E3) which were more pronounced during contralateral than during ipsilateral activations.

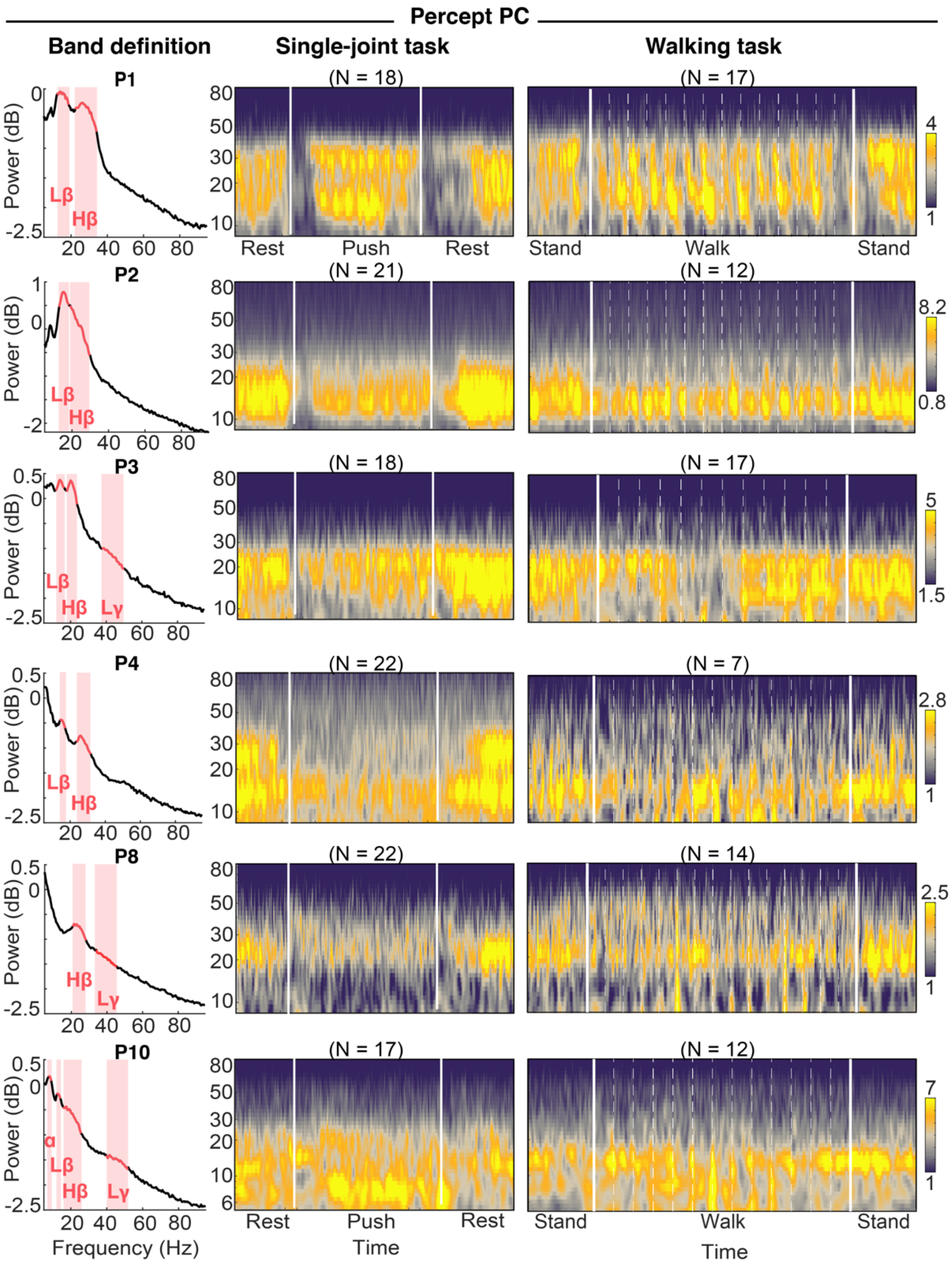


**Figure S5 | Idiosyncratic encoding of leg muscle activation across tasks for participants implanted with a Percept PC stimulator.** Left column: Power spectral densities highlight individual frequency bands across patients, as identified using an unbiased fitting algorithm (Donoghue, T. *et al.*). Middle column: Average (not normalized) scalograms during an isometric knee extension task in the neurorobotic platform. Vertical lines indicate the start and release of muscle contraction. Right column: Average scalograms (not normalized) for the walking task. Vertical lines indicate the initiation and termination of walking. Dashed lines correspond to foot strike events.

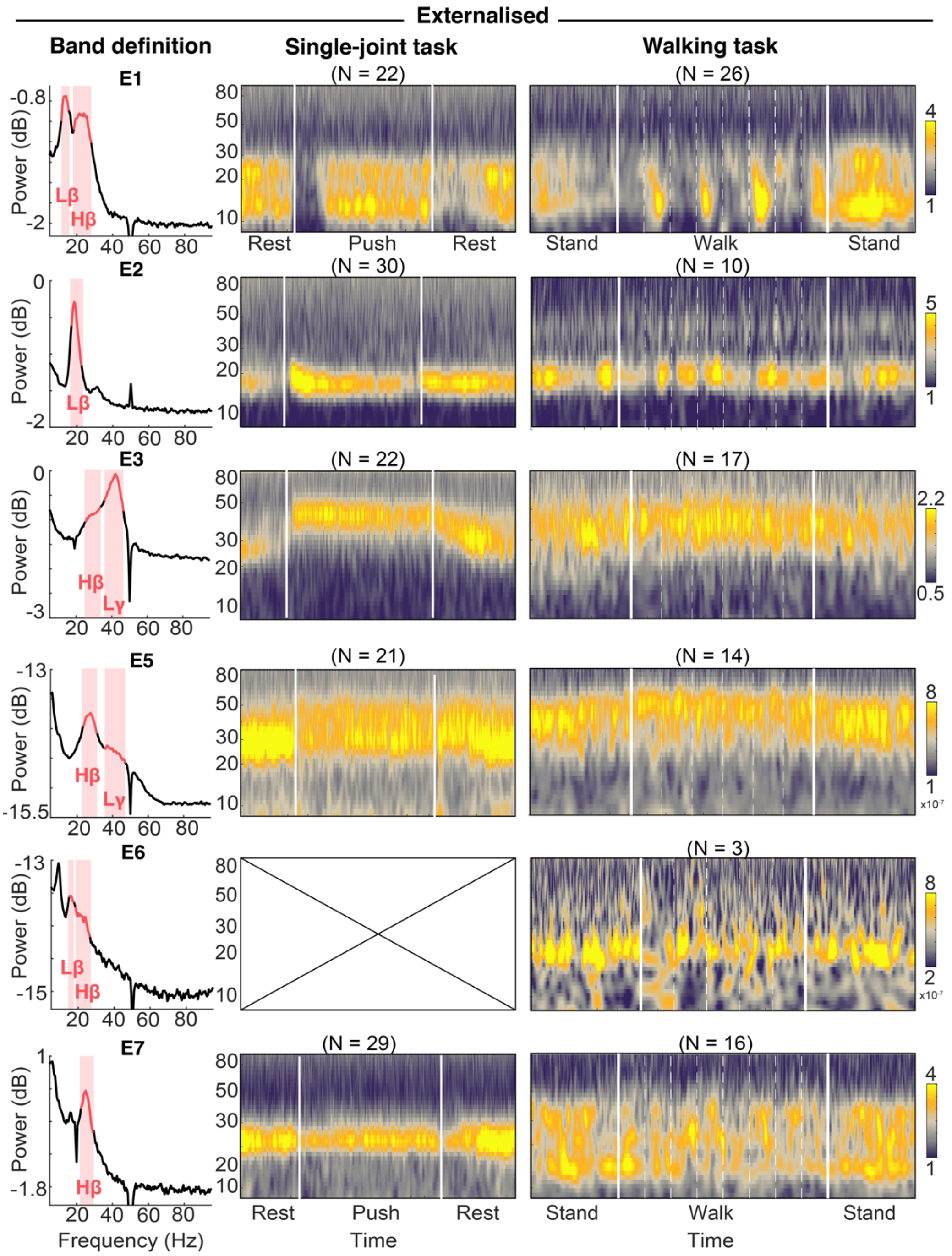


**Figure S6 | Idiosyncratic encoding of leg muscle activation across tasks for participants recorded while their DBS leads externalized.** Same as Extended Data Figure 5.

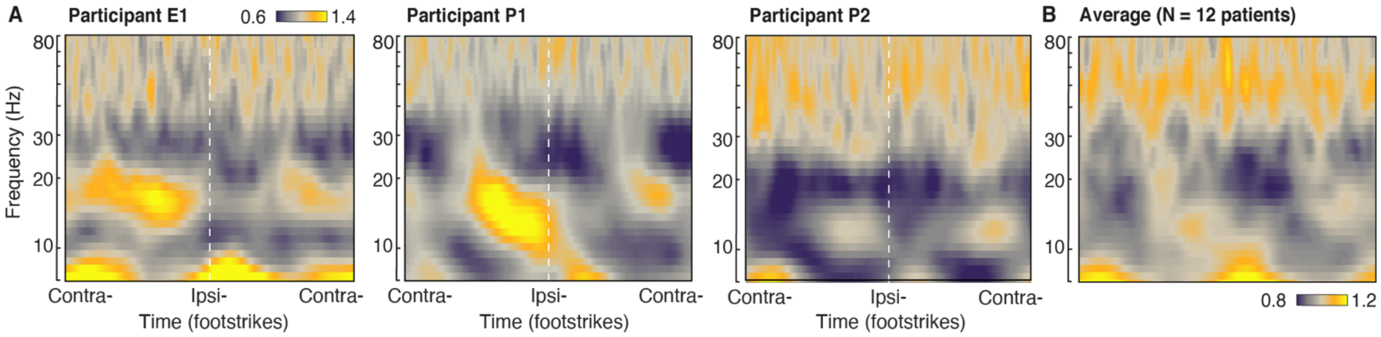


**Figure S7 I STN LFP modulations per gait-cycle. (A)** Illustrative examples of gait-cycle averaged spectrograms for three participants (one externalized E1 and two Percept P1 and P2), highlighting similar temporal patterns, yet idiosyncratic frequency bands and modulation amplitudes. **(B)** Cross-patient average (N = 12).

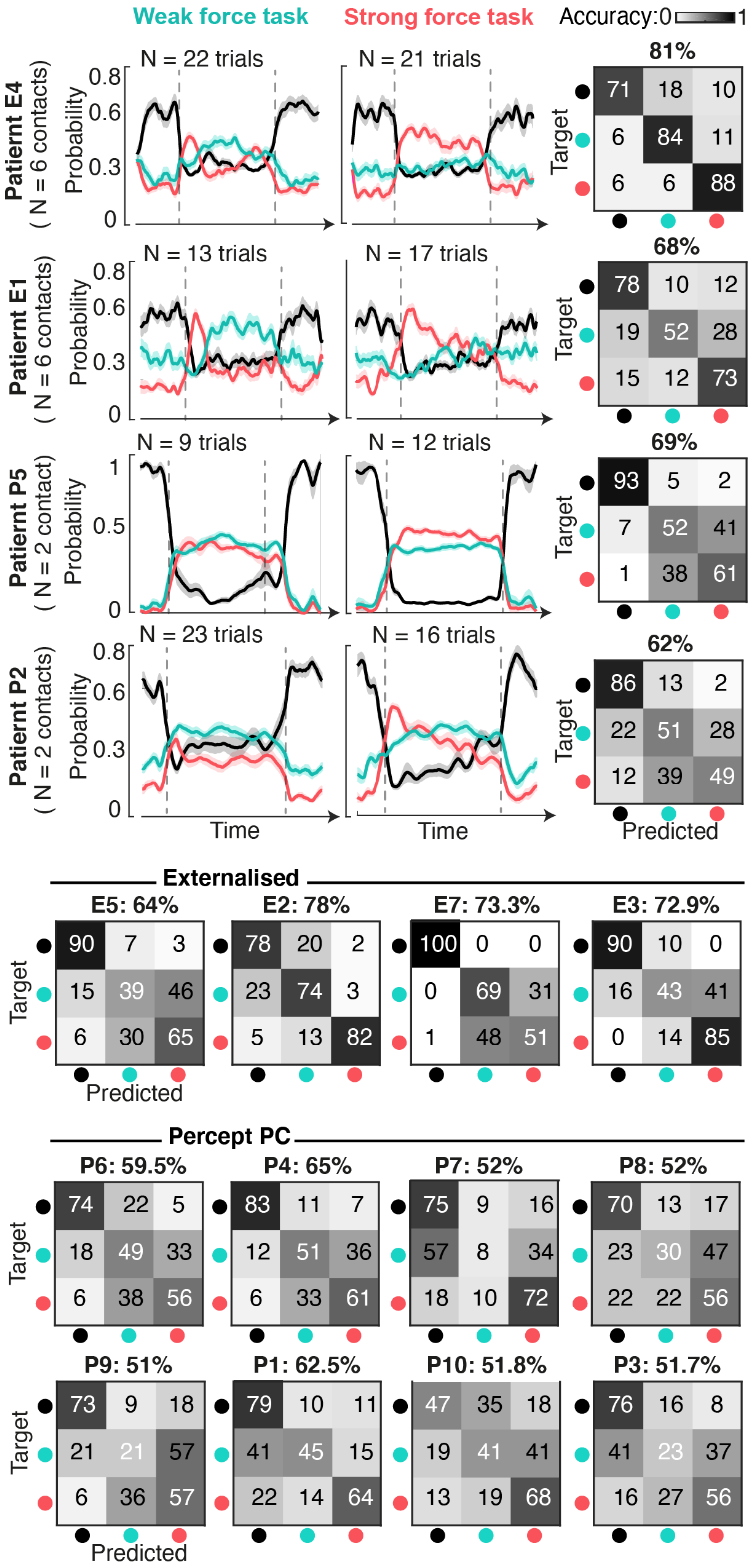


**Figure S8 I Decoding performance of single-joint vigor tasks across participants.** Temporal profiles of probability traces for four illustrative participants (two externalized E1 and E4, and two Percept P2 and P5), separately for weak and strong forces, along with sample-based confusion matrices across tasks of all the patients.

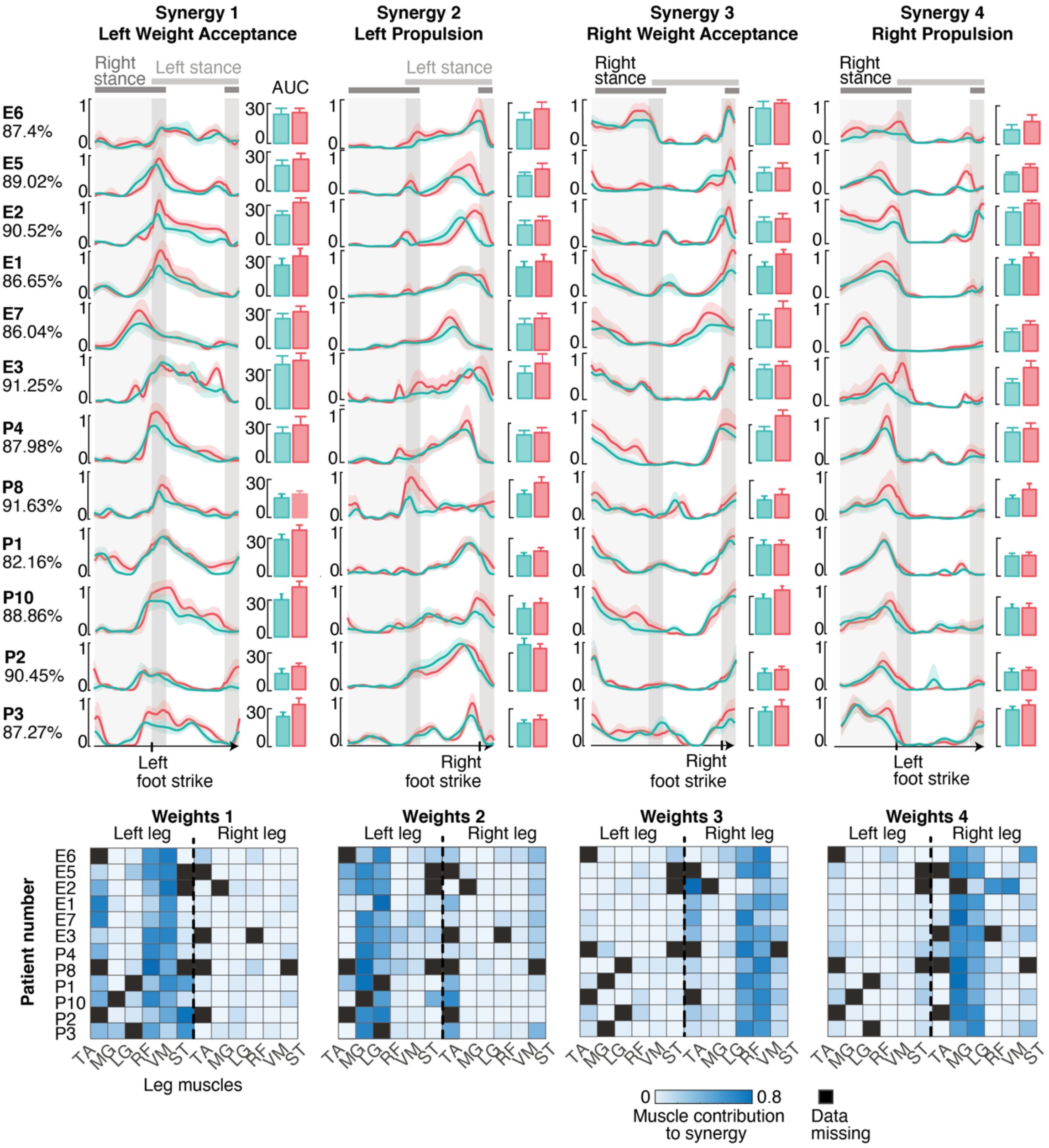


**Figure S9 I Muscle synergies across participants.** Temporal profiles of all extracted leg muscle synergies, percentage of variance explained and weights of muscle contribution for all the individual participants who performed the walking tasks.

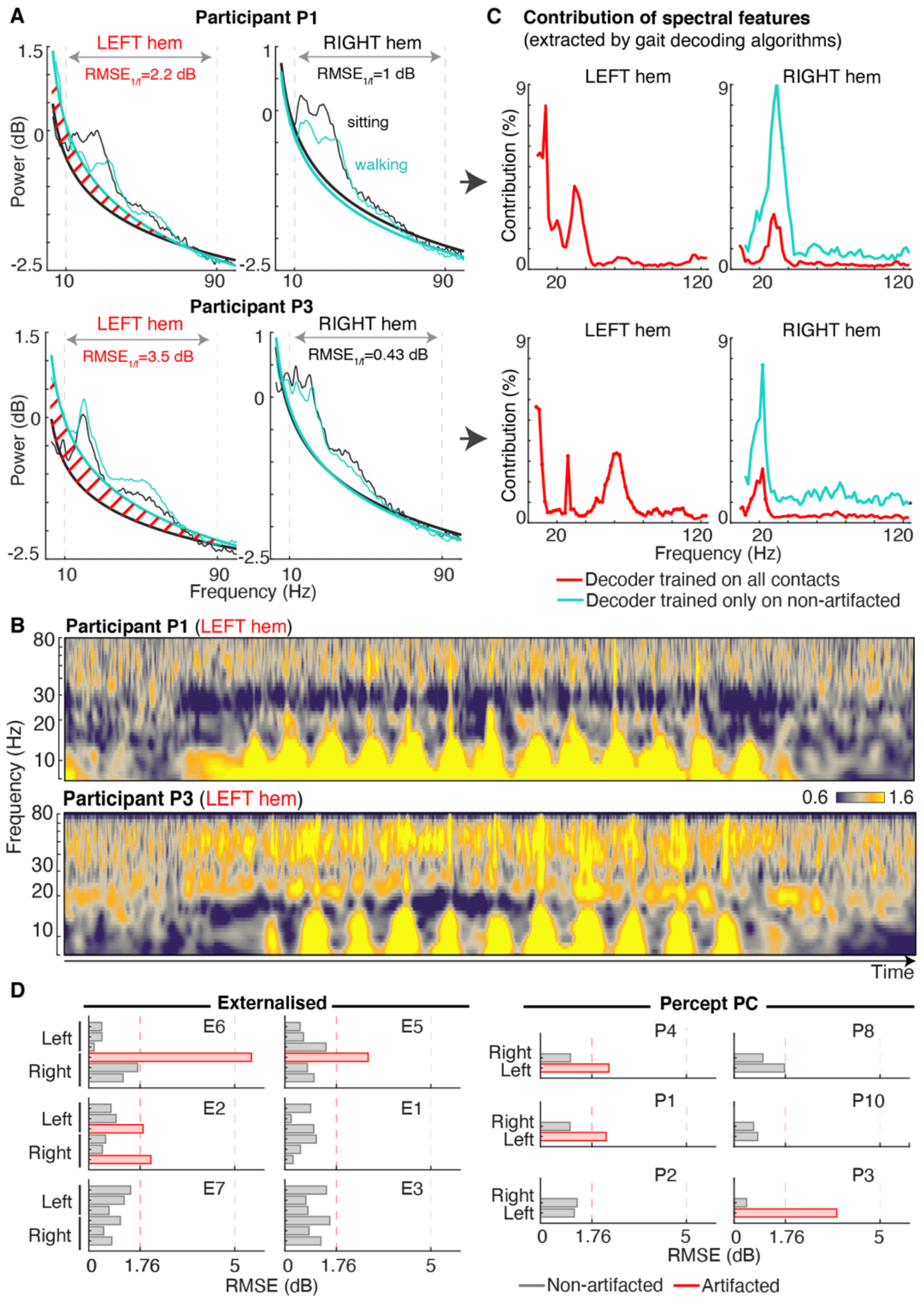


**Figure S10 I Identification of artefacted channels during walking.** For each contact pair, we compared the aperiodic (1/f) components of the PSD computed during sitting versus walking. Contacts that exhibited an increase in energy of 50% or more (1.76 dB) during walking in the range of interest (10-90Hz) were labeled as artefacted. **(A)** Illustrative example of bilateral PSD for two participants (P1 and P3), for whom one hemisphere was artefacted, and **(B)** spectrogram of the corrupted channels, showing gait-locked artifacts that emerge from low frequencies and propagate to high frequencies. **(C)** Comparison of the spectral features that contribute to the decoding algorithms when trained using all contacts (red) as compared to only using non-artefacted contacts (green). Artefacted channels (Left hemisphere for P1 and P3) systematically exhibit predominant contributions of very low frequencies. Their removal does not significantly change the frequencies contributing from other channels. **(D)** Identification of artefacted channels for all contacts and patients.

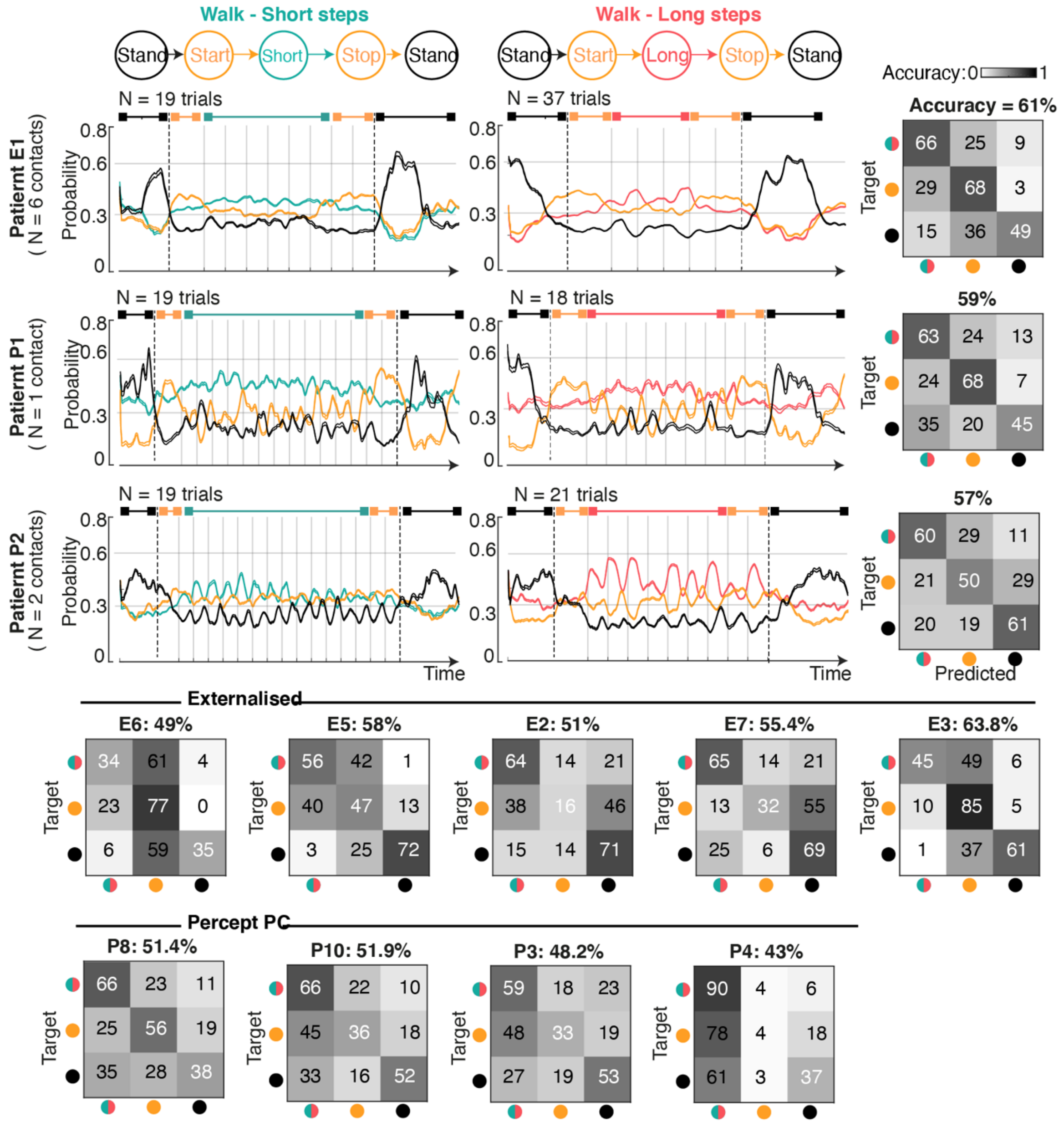


**Figure S11 l Decoder of walking states across participants.** Temporal profiles of probability traces for three participants (one externalized E1 and two Percept P1 and P2), separately for short and long steps, and sample-based confusion matrices of all participants.

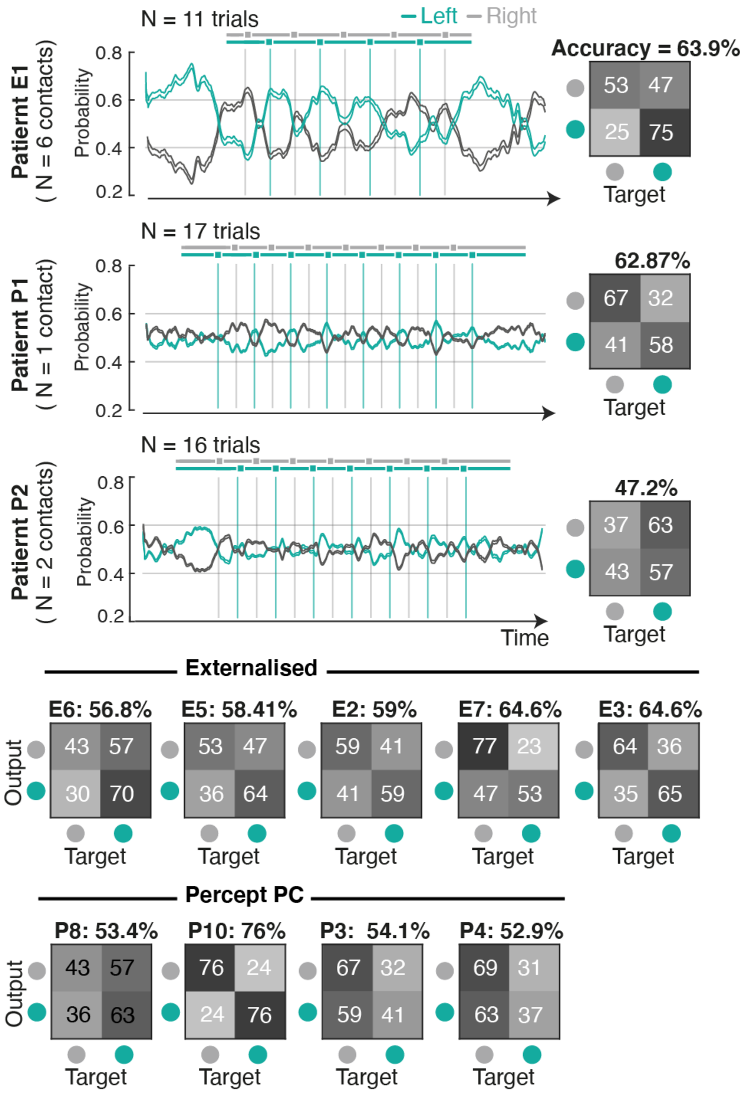


**Figure S12 l Decoder of gait events across participants.** Temporal profiles of probability traces for three participants (same as in Extended Data Fig. 10), along with sample-based confusion matrices of all the participants.

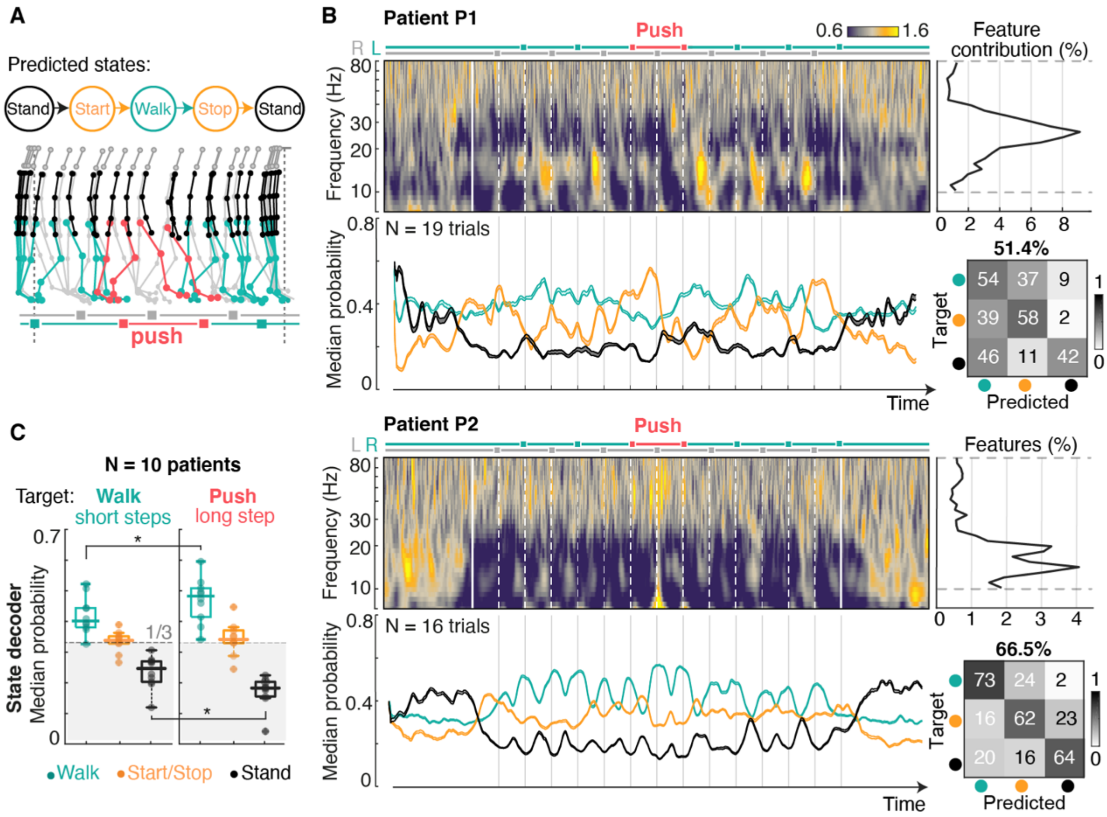


**Figure S13 | Performance of walking-state decoder during gait adaptations. (A)** We tested the walking-state decoder, trained on short and long steps, during a task requiring a sudden increase in effort during walking. **(B)** Illustrative examples of average spectrograms and probability traces for two patients. The step requiring a sudden increase in vigor (push) was either decoded as a transition (patient P1) due to the similarities to initiation and termination patterns, or with a higher probability of walking (patient P2). This depended on the amplitude of modulations in each individual patient. **(C)** Across all patients, the step with increased vigor translated into an increase of walking probabilities and a decrease of rest probabilities.

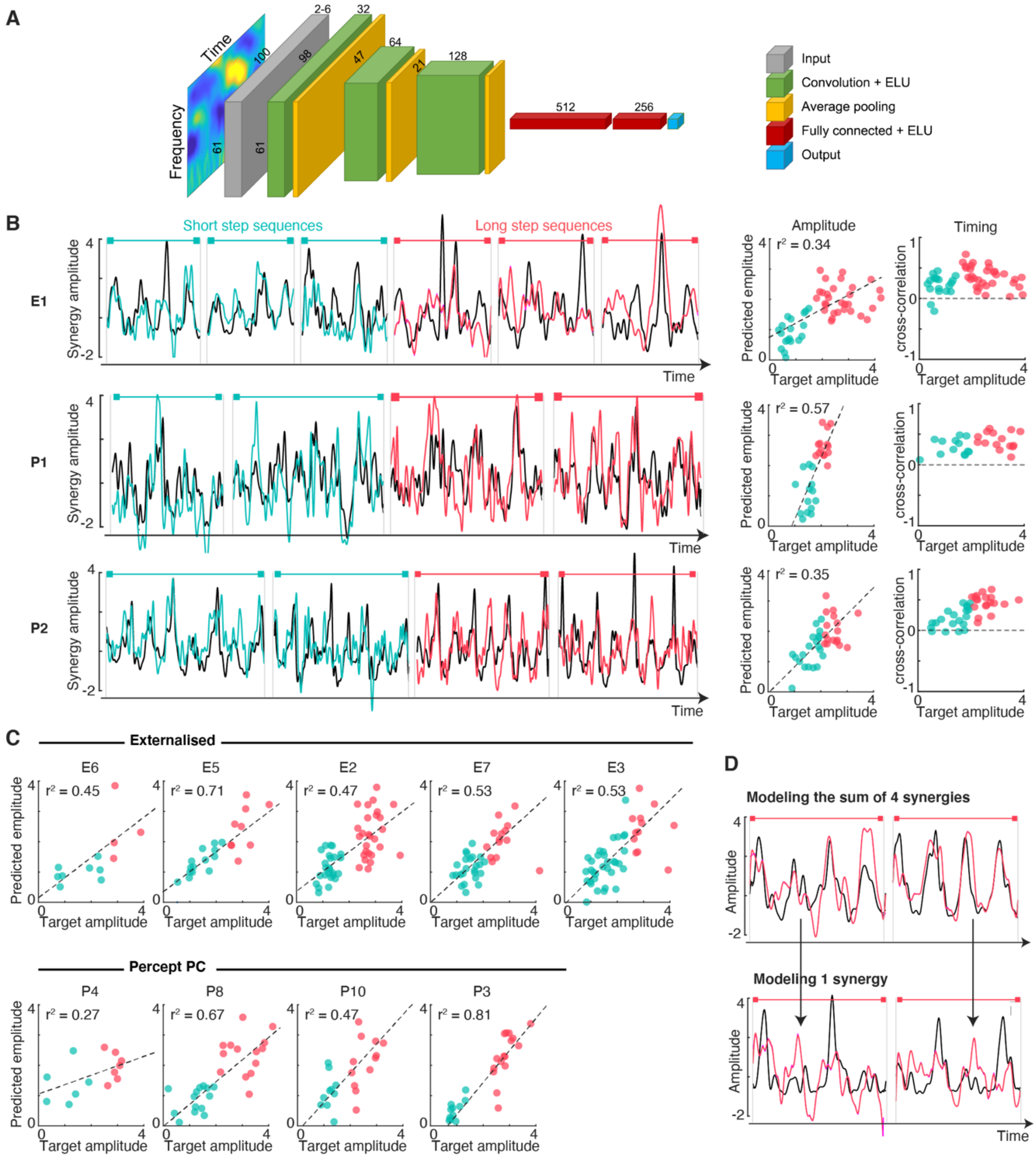


**Figure S14 | Model of leg muscle synergy profiles using a deep learning algorithm across participants. (A)** Structure of the deep neural network. **(B)** Comparison of target and modeled synergy traces for one externalized and two Percept PC patients (same as in Extended Data Fig. 10), and **(C)** performance across patients. **(D)** Illustrative example of prediction output when modeling either all bilateral synergies (top) versus only one unilateral synergy (bottom). Using STN LFP, the deep learning model automatically predicted modulations happening twice per gait cycle, as expected from the bilateral encoding of leg movements in the STN.

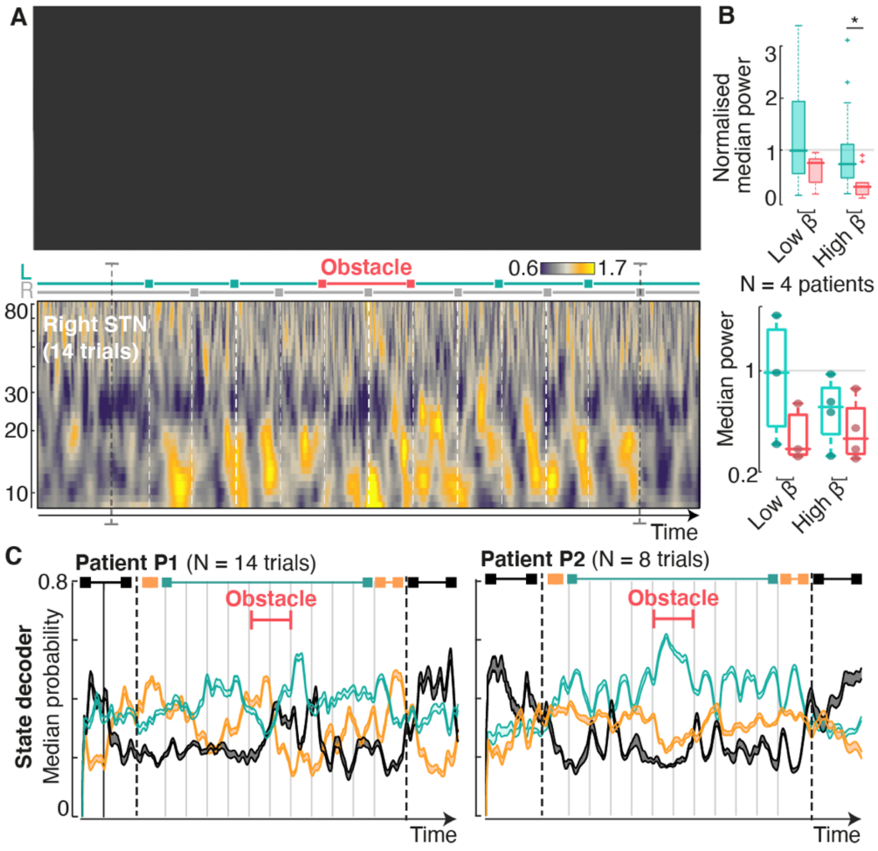


**Figure S15 l STN LFP modulations during activities of daily living. (A)** Time decomposition of a walking sequence (participant P1) that involves stepping over an obstacle. Chronophotography (top) and average spectrogram (bottom) highlight modulations in low- and high-beta that mirror the patterns observed during the horizontal ladder tasks. **(B)** Avoiding the obstacle induced modulations in low- and high-beta power across all tested patients (N=4). **(C)** Similarly to what was observed during the sudden long step, the decoder (trained on short and long steps) predicted the obstacle as either a transition or with a bigger probability of walking (compared to rest).

| ID | Sex | Age range | Disease duration (years) | Dominant symptoms (AR or TD) | LEDD pre-op (mg eq.) | Total UPDRSIII (OFF/ON med, pre-Sx) | Most affected hemibody | Setup | Single-joint movement | Walk | Medication status |
|---|---|---|---|---|---|---|---|---|---|---|---|
| **E4** | M | 60-69 | 12 | AR+T | 1437.5 | 27/14 (UPDRS) | Right | Externalized | X | | OFF > 12h |
| **E6** | M | 60-69 | 7 | AR | 1066,7 | 23/11 (UPDRS) | | Externalized | | X | OFF > 12h |
| | | | | | | | | | | X | Residual |
| **E5** | M | 50-59 | 13 | AR | 1453,3 | 25/12 (MDS-UPDRS) | Right | Externalized | X | X | Residual |
| **E2** | M | 50-59 | 7 | AR | 700 | 30/7 (/142) | Right | Externalized | X | X | OFF > 12h |
| **E1** | M | 70-79 | 6 | AR+T | 1087,5 | 43/23 (UPDRS) | Right | Externalized | | X | OFF > 12h |
| | | | | | | | | | X | X | Residual |
| **E7** | M | 60-69 | 15 | AR | 1600 | 35/14 | Right | Externalized | | X | Residual |
| **E3** | M | 60-69 | 11 | AR+T | 1150 | 37/18 (MDS-UPDRS) | Left | Externalized | X | X | Residual |
| **P6** | M | 70-79 | 8 | AR | 1670 | 31/9 (MDS-UPDRS) | Right | Percept | X | X | Residual |
| **E8** | F | 40-49 | 5 | AR+T | 500 | 63/12 (/108) | Left | Externalized | X | X | ON |
| **P4** | M | 60-69 | 7 | AR+T | 1350 | 45/33(1h)/18(2h) (MDS-UPDRS) | Right | Percept | X | X | ON |
| **P5** | M | 70-79 | 13 | AR | 1600 | 30/9 (MDS-UPDRS) | Left | Percept | X | | ON |
| | | | | | | | | | X | | OFF > 12h |
| **P7** | M | 50-59 | 7 | AR+T | 475 | 50/30 (MDS-UPDRS) | Right | Percept | X | | ON |
| | | | | | | | | | X | | Residual |
| **P8** | M | 60-69 | 14 | AR+T | 1325 | 26/8 | Right | Percept | X | | OFF > 12h |
| **P9** | M | 60-69 | 11 | AR+T | 1150 | 43/19 (MDS-UPDRS) | Right | Percept | X | | OFF > 12h |
| | | | | | | | | | X | X | |
| **P1** | F | 50-59 | 9 | AR+T | 1050 | 17/6 (MDS-UPDRS) | Right | Percept | X | | OFF > 12h |
| | | | | | | | | | | X | Residual |
| **P10** | M | 50-59 | 17 | T | 1665 | 46/12 (MDS-UPDRS) | Right | Percept | | X | |
| | | | | | | | | | X | | Residual |
| **P2** | M | 70-79 | 8 | AR+T | 1300 | 44/16 | Right | Percept | X | X | OFF > 12h |
| **P3** | M | 60-69 | 11 | AR+T | 1050 | 38/18 MDS-UPDRS | Left | Percept | X | X | ON |

**Table S1 | Clinical details for all patients**. Participants whose complete Parkinsonian medication was withdrawn 12h prior to experiments are indicated as "OFF>12h". Participants who received some Levodopa medication within 3h before the experiment are labeled "ON". Others (e.g., participants who retained some agonist medication, or those who received a delayed Levodopa dose the night before the experiment) are labeled as having some "residual" medication.
